# Metabolome-wide association of carotid intima media thickness identifies FDX1 as a determinant of cholesterol metabolism and cardiovascular risk in Asian populations

**DOI:** 10.1101/2024.05.14.24307316

**Authors:** Nilanjana Sadhu, Rinkoo Dalan, Pritesh R Jain, Chang Jie Mick Lee, Leroy Sivappiragasam Pakkiri, Kai Yi Tay, Theresia H Mina, Dorrain Low, Yilin Min, Matthew Ackers-Johnson, Thi Tun Thi, Vishnu Goutham Kota, Yu Shi, Yan Liu, Henry Yu, Darwin Tay, Hong Kiat Ng, Xiaoyan Wang, Kari E Wong, Max Lam, Xue Li Guan, Nicolas Bertin, Eleanor Wong, James Best, Rangaprasad Sarangarajan, Paul Elliott, Elio Riboli, Jimmy Lee, Eng Sing Lee, Joanne Ngeow, Patrick Tan, Christine Cheung, Chester Lee Drum, Roger SY Foo, Gregory A Michelotti, Haojie Yu, Patricia A Sheridan, Marie Loh, John C Chambers

## Abstract

The burden of cardiovascular disease (CVD) is rising in the Asia-Pacific region, in contrast to falling CVD mortality rates in Europe and North America. To provide new insights into the pathways influencing cardiovascular risk in Asian populations, we quantified 883 metabolites by untargeted mass spectroscopy in 8,124 Singaporean adults, and investigated their relationships to carotid intima media thickness (cIMT), a marker of atherosclerosis. We found that plasma concentrations of 3beta-hydroxy-5-cholestenoate (3BH5C), a cholesterol metabolite, associated inversely with cIMT (Beta[SE]=-0.013[0.002]). Genetic instruments support a causal relationship of 3BH5C metabolic pathways on cardiovascular risk, with a 5-6 fold higher effect size in Asians (OR_GSMR_[95% CI]=0.89[0.87-0.92], OR_IVW_[95% CI]=0.86[0.80-0.92]) compared to Europeans (OR_IVW_[95% CI] = 0.98[0.96-0.99]). Colocalization analyses indicate the presence of a shared causal variant between 3BH5C plasma levels and expression of ferredoxin-1 (FDX1), a protein essential for sterol and bile acid synthesis. We validated *FDX1* as a key regulator of 3BH5C synthesis in hepatocytes, and macrophages, and cholesterol efflux in aortic smooth muscle cells, through knockout and overexpression models. In summary, this study makes an important contribution to our understanding of the metabolic basis for atherosclerosis in Asian populations, and identifies *FDX1* as a potential therapeutic target for prevention of CVD.

## Introduction

Atherosclerosis is a chronic inflammatory vascular disease characterized by the accumulation of cholesterol and other lipids, immune cells, and fibrotic tissue in arterial walls leading to the formation of atherosclerotic plaques^1^. The progressive narrowing and hardening of arteries, coupled with superadded thrombosis, can cause a range of cardiovascular complications, including myocardial infarction, stroke, heart failure and critical limb ischemia. Atherosclerosis remains a major global health problem, as the primary contributor to cardiovascular disease (CVD) worldwide^2^.

Globally, CVD is the leading cause of mortality^3^. However, there is increasing recognition that there are important differences in CVD development and progression between populations^4^. In contrast to the stable or falling incidence of CVD events in North America and Europe, the risk of fatal and non-fatal CVD is high and rising in Asian populations ^5,6^. Asia already accounts for greater than 50% of global CVD deaths, and the proportion of premature CVD deaths in Asia (∼39%) is higher than that of Europe or the US (22-23%)^7^. The reasons underlying these divergent outcomes for CVD between populations are not well understood, but are likely to reflect the presence of genetic and environmental factors that are specific to or more prevalent in Asian populations. Understanding how atherosclerosis develops and manifests in Asian populations may help identify novel targets for CVD prevention and treatment, that may be of wider global relevance.

Metabolites are a reflection of cellular processes involved in complex biological pathways. Since the metabolome lies downstream of all other molecular processes and is proximal to clinical phenotypes, metabolites can serve as potential endpoint biomarkers of diseases^8,9^. Advanced analytical techniques have enabled exploration of the metabolome in an efficient and standardized manner^10,11^. Untargeted profiling of circulating metabolites in a population cohort, thus, provides an opportunity to identify metabolic patterns and pathways specific to disease conditions such as atherosclerosis. Such studies can also facilitate comparison of metabolic signatures of diseases across different populations.

We therefore performed untargeted metabolic profiling and assessment of cardiovascular health, amongst 8,124 Asian men and women participating in a well characterized longitudinal population cohort, to identify novel mechanisms underlying atherosclerosis and CVD.

## Results

### Metabolic variation and Carotid Intima Media Thickness

Our experimental design is summarized in **Fig. 1**. We first performed a metabolome-wide association study of carotid intima media thickness (cIMT), a validated marker of cardiovascular risk^12^, amongst 8,124 Asian participants from the Health for Life in Singapore (HELIOS) study. The characteristics of participants are summarized in **Supplementary Table 1**. Median age of cohort participants was 54 years, and 59% were female. The ethnic composition was Chinese 67%, Malay 14%, and Indian 19%. Mean±SD body mass index (BMI) was 25±5 kg/m^2^ and 82% reported as never smokers. The prevalence of obesity (BMI ≥ 30kg/m^2^), diabetes, and hypertension was 12%, 12%, and 29%, respectively. cIMT was measured in the common carotid artery using a Philips iU 22 ultrasound system; mean cIMT of participants was 0.60±0.13 mm (**Supplementary Fig. 1**).

**Figure 1.**
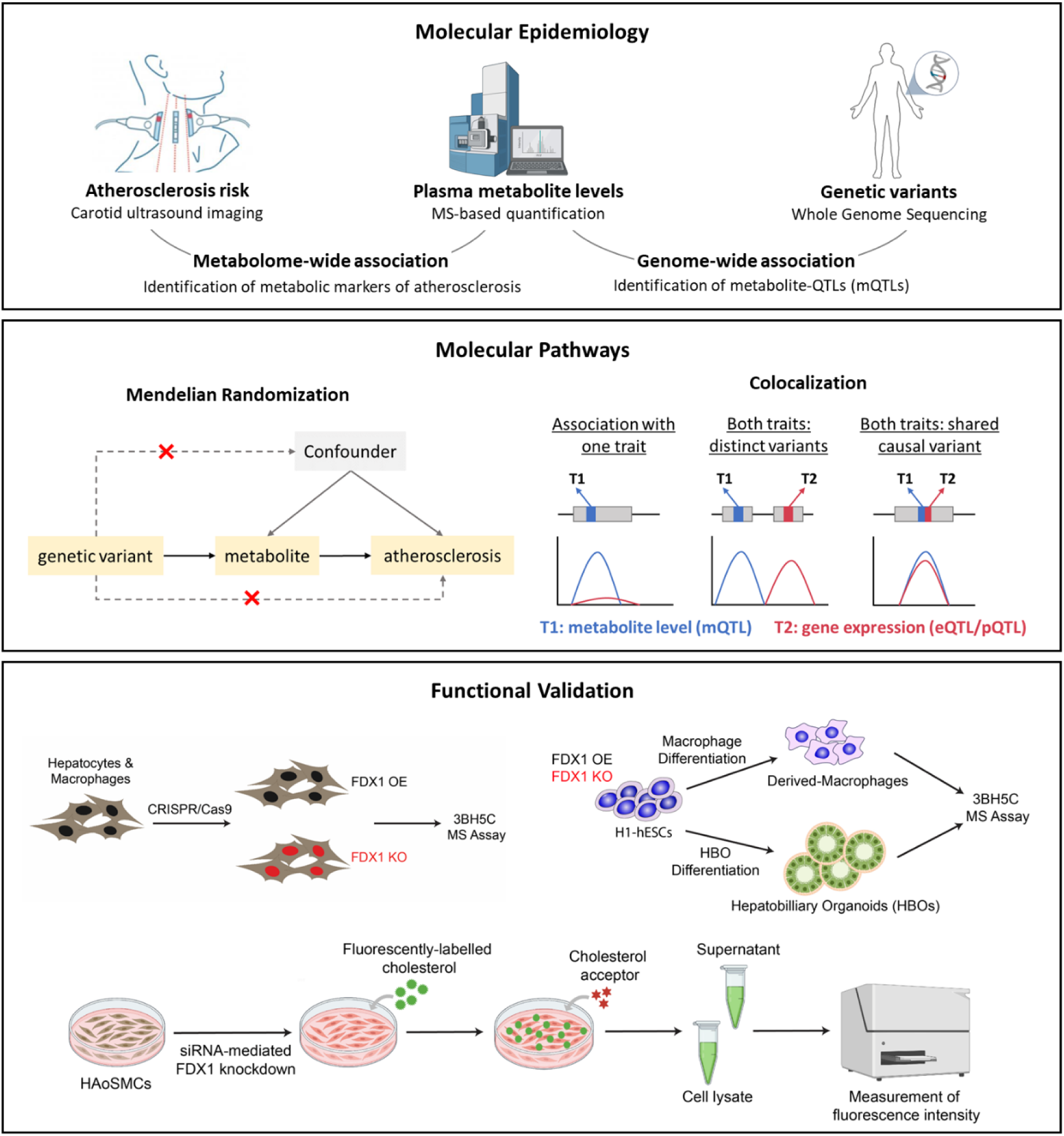
Summary of Study Design. 3BH5C, 3beta-hydroxy-5-cholestenoate; eQTL, expression Quantitative Trait Loci; HAoSMCs, Human Aortic Smooth Muscle Cells; H1-hESCs, H1-derived Human Embryonic Stem Cells; HBOs, Hepatobiliary Organoids; FDX1, Ferredoxin-1; KO, Knock-out; MS, Mass Spectroscopy; mQTL, metabolite Quantitative Trait Loci; OE, Over-expression; pQTL, protein Quantitative Trait Loci. Image partially created using Biorender.com

883 metabolites were quantified in fasting plasma samples using multimodal mass spectrometry (Metabolon Global Discovery panel, **Supplementary Tables 2-3**). The majority of these metabolites were lipid species (40%), amino acids (19.5%) or xenobiotics (10%). Approximately 18% of the metabolites were partially characterized or X-compounds (**Supplementary Fig. 2**).

Linear regression revealed that 252 metabolites, including 33 X-compounds, associated with cIMT after adjusting for age, sex, ethnicity, and batch (P<5.7×10^-5^ accounting for Bonferroni-correction, **Supplementary Table 4**). Amongst these metabolites, 126 (∼50%) remained associated with cIMT (P<0.05, and same direction of effect), after accounting for traditional CV risk factors, including BMI, systolic blood pressure (BP), total cholesterol, Type-2 Diabetes (T2D) status, and smoking status (**Supplementary Table 5**).

### Causal inference testing through Mendelian randomization

We carried out Mendelian randomization (MR) analysis, to identify potential causal relationships between each of the 126 metabolites as exposures and two outcomes: (i) cIMT as a quantitative measure of vascular risk, and (ii) coronary artery disease (CAD) as the major clinical consequence of atherosclerosis. We first applied the generalized summary-based Mendelian randomization (GSMR) method^13^ on genome wide association study (GWAS) summary statistics from (i) HELIOS for each metabolite exposure, (ii) UK Biobank (UKBB) for the cIMT outcome^14^, and (iii) Biobank Japan (BBJ) for the CAD outcome^15^. Independent genetic instruments for each metabolite exposure were selected at the suggestive genome-wide significant threshold of P <1×10^-5^, and clumping performed at r^2^ threshold = 0.05, window size = 1mb. Genetic variants with pleiotropic effects (on both exposure and outcome) were detected using the HEIDI-outlier approach^13^ (P <0.01) and removed. Amongst the 126 metabolites tested, two lipid metabolites: 3beta-hydroxy-5-cholestenoate (3BH5C, P = 1.5×10^-16^), and 1-linoleoyl-2-arachidonoyl-GPC (18:2/20:4n6)* (P = 1.4×10^-4^) showed evidence for having a potential causal relationship with CAD at P < 4×10^-4^ accounting for multiple testing (**Supplementary Table 7**). In contrast, none of the 126 metabolites showed evidence that might indicate a direct causal relationship with cIMT (**Supplementary Table 6;** all Bonferroni corrected P>0.05).

We further evaluated the relationship between these two metabolites and CAD by TwoSampleMR^16^, using meta-analyzed GWAS summary data from independent predominantly-European cohorts (metabolite exposures: CLSA^17^, METSIM^18^; outcome CAD: UKBB^19^, CARDIoGRAMplusC4D^20^). We applied different MR methods (inverse-variance weighted [IVW], weighted-median, weighted-mode), and demonstrated that the protective effect of 3BH5C on CAD was robust, with a consistent direction of effect (**Fig. 2**, **Supplementary Table 8**). There was no evidence for reverse causality or for horizontal pleiotropy (P_egger_intercept_ = 0.48, P_MR-PRESSO_Global-Test_ > 0.05 [no outliers detected]). The causal estimate was also robust to the choice of genetic instruments based on IVW leave-one-out sensitivity analysis (**Supplementary Fig. 3**).

**Figure 2.**
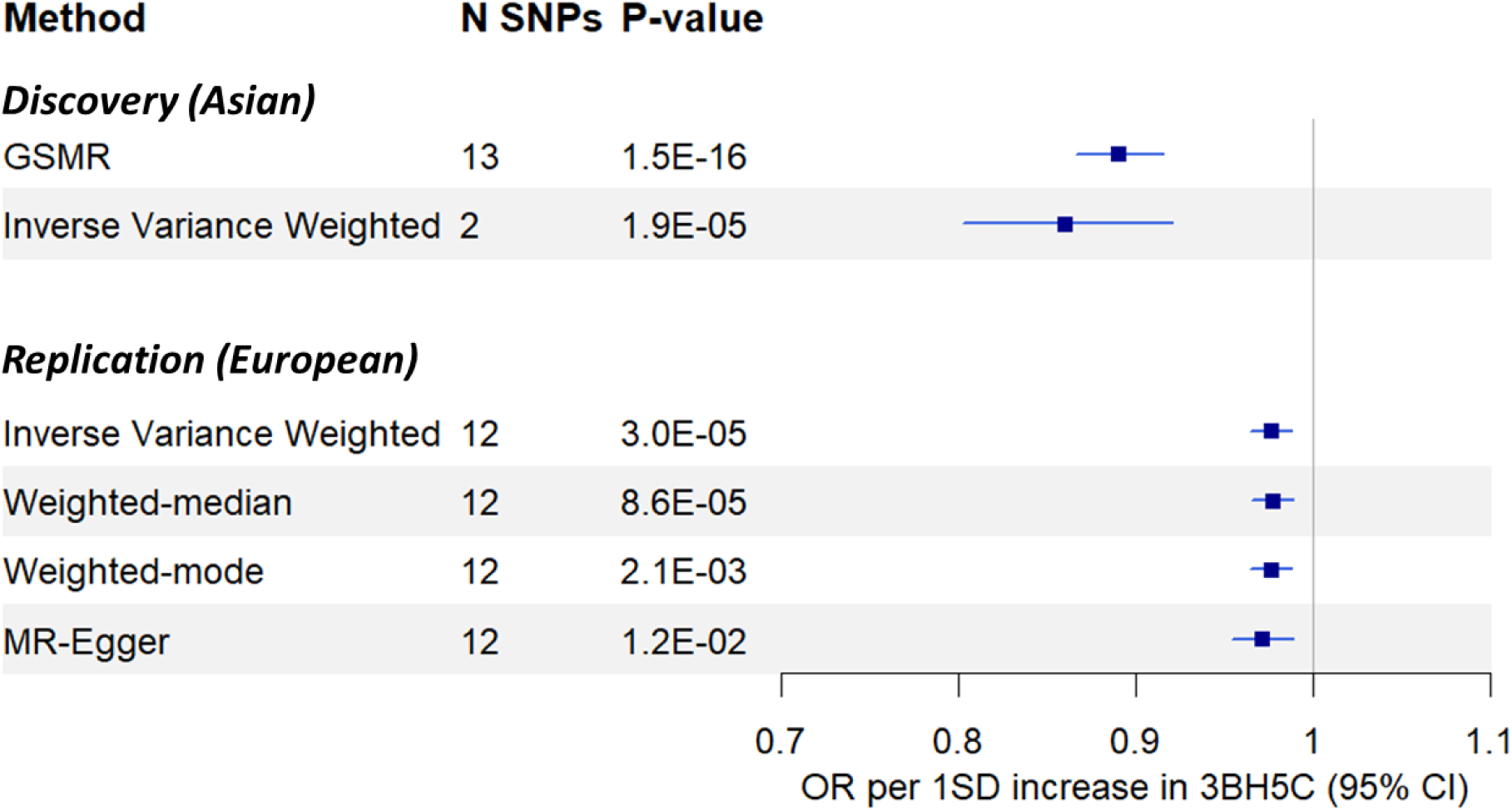
Summary of results from Mendelian randomization studies evaluating effect of exposure 3BH5C levels on outcome CAD risk. Discovery analysis using GWAS summary data from HELIOS for exposure 3BH5C, and BBJ for outcome CAD. Replication analysis using meta-analyzed GWAS summary data from CLAS and METSIM for exposure 3BH5C, and UKBB and CARDIoGRAMplusC4D for outcome CAD. Instrument selection criteria for GSMR: p<1e-05, r²<0.05, 1000G, HEIDI outlier filtering p<0.01. Instrument selection criteria for TwoSampleMR methods (Inverse Variance Weighted, Weighted-median, Weighted-mode, MR-Egger): p < 5×10^-8^, r^2^ = 0.05, window = 1000kb, 1000G, F-statistic >=30. Test for heterogeneity between Discovery IVW and Replication IVW: P_heterogeneity_ < 0.0001, I^2^ = 94.07%. 3BH5C, 3beta-hydroxy-5-cholestenoate; CAD, Coronary Artery Disease; GSMR, Generalized Summary-based Mendelian Randomization

Our MR analyses support a protective effect of 3BH5C metabolic pathways on CAD, in both Asian and European populations, with a ∼5-6-fold higher effect size in the former (odds ratio for CAD per 1SD increase in plasma 3BH5C concentrations: Asian population OR_GSMR_ [95% CI] = 0.89 [0.87-0.92], P_GSMR_ = 1.5×10^-16^; Asian population OR_IVW_ [95% CI] = 0.86 [0.80-0.92], P_IVW_ = 1.9×10^-5^; European populations OR_IVW_ [95% CI] = 0.98 [0.96-0.99], P_IVW_ = 3.0×10^-5^; heterogeneity test between Asian and European IVW-based MR effect estimates: P_het_ = 0.0003, I^2^ = 92.18%, **Supplementary Table 8**). Concentrations of plasma 3HB5C were lowest in Indian and Malay individuals (P = 8.7×10^-4^; **Supplementary Table 3**), consistent with previously reported higher risk of CVD in these Asian ethnic sub-groups, compared to individuals of Chinese ethnicity^21,22^. There was also no evidence for heterogeneity of effect on the relationship between 3BH5C and cIMT, between the Asian subgroups in our study cohort (P_heterogeneity_ = 0.2; **Supplementary Table 4**).

The effect of 1-linoleoyl-2-arachidonoyl-GPC (18:2/20:4n6)* on CAD in Asian populations(OR [95% CI] = 0.84 [0.75-0.93], P_Wald-ratio_ = 1.1×10^-3^) is driven by a genetic instrument in *FADS1* – a well-known and highly pleiotropic CVD-risk locus^23^. However, the relationship of this metabolite with CVD was not significant in MR analysis using independent European cohorts (P_IVW_ = 0.50, **Supplementary Table 8**).

### Genetic regulation of 3BH5C and CVD risk

Genome-wide association in the HELIOS study revealed 404 variants to be associated with circulating 3BH5C levels at P<5×10^-8^. These are located in a single region on chromosome 11, between genes *RDX* and *FDX1*, with the top variant being rs2051466 (Beta [SE] per copy of the A allele = 0.50 [0.04], P = 7.9×10^-43^, λ_GC_ = 1.002; **Supplementary Fig. 4**). Our results are replicated by publicly available data from the METSIM (N=6,136) and CLSA (N=8,299) studies, which confirm the strong association of rs2051466 with 3BH5C concentrations (Beta_METSIM_ [SE] = 0.39 [0.02], P_METSIM_ = 4.4×10^-90^; Beta_CLSA_ [SE] = 0.31 [0.02], P_METSIM_ = 1.2×10^-74^).

FUMA-based functional annotation^24^ of 1,259 candidate genetic variants in this locus (P<0.05 and r^2^≥0.05 with top SNP rs2051466, see **Methods**) showed that 80% of the variants were enriched in regions of transcriptional/enhancer activity. 56% of the candidate variants were intergenic, and 1.6% had a RegulomeDB score <2 providing evidence for the variants being likely to have a regulatory function (**Supplementary Table 9**). Fine-mapping of a 3mb region around rs2051466 for association with 3BH5C using GWAS data from HELIOS identified rs10488763 (PIP = 0.50) and rs1443120 (PIP = 0.50) as potentially causal variants, based on 95% credible sets. These genetic variants are neither coding variants, nor in strong LD (r^2^>0.8) with a coding variant, further arguing for regulatory disturbances, rather than an effect on protein structure, as the genetic mechanism.

Both genetic variants, rs10488763 and rs1443120, are reported to be associated with CAD in the Japanese population^15^ (**Fig. 3**), as well as in trans-ethnic meta-analyses of Japanese and European populations^15,25^. However, their associations with CAD are not genome-wide significant in predominantly European populations^19,20^(**Fig. 3**). The two genetic variants are in strong linkage disequilibrium (LD r^2^ = 0.99). In keeping with this observation, the strength, size, and direction of effect of rs10488763 and rs1443120 on 3BH5C and CAD are highly comparable ^25^. The direction of effect indicates that a decreased 3BH5C concentration is associated with an increased risk of CAD (associations summarized in **Fig. 3**). rs10488763 is also associated with myocardial infarction, stable angina pectoris, and use of vasodilators, salicylic acid, and antithrombotic agents within Biobank Japan (Bonferroni-corrected P<0.05; **Supplementary Fig. 6**).

**Figure 3.**
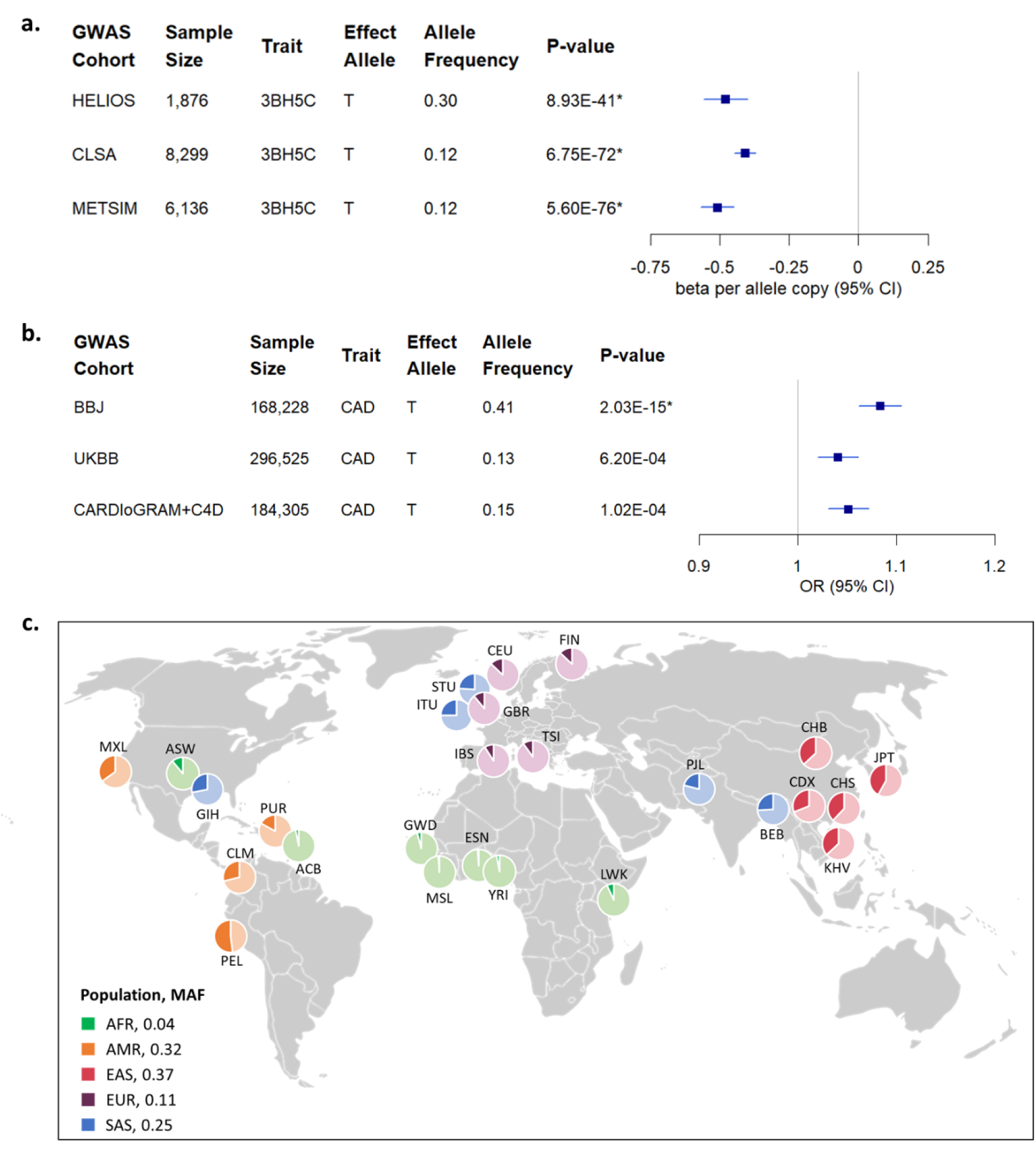
Association of rs10488763 with 3BH5C and CAD in different populations, and its global variation in allele frequency. (a) Forest plot summarising the association of rs10488763 with 3BH5C and (b) with CAD from GWAS in different cohorts. *indicates genome-wide significance (P<5×10^-8^). 3BH5C, 3beta-hydroxy-5-cholestenoate; CAD, Coronary Artery Disease; CLSA, Canadian Longitudinal Study on Aging; METSIM, Metabolic Syndrome in Men study; BBJ, Biobank Japan; UKBB, UK Biobank. (c) Global distribution of minor allele frequency (MAF) of rs10488763 (minor allele T) based on 1000 Genomes Project. AFR, African; AMR, American; EAS, East Asian, EUR, European, SAS, South Asian. ACB, African Caribbean in Barbados; ASW, African Ancestry in SouthWest US; ESN, Esan in Nigeria; GWD, Gambian in Western Division; LWK, Luhya in Webuye Kenya; MSL, Mende in Sierra Leone; YRI, Yoruba in Ibadan. CLM, Colombian in Medellin; MXL, Mexican Ancestry in Los Angeles; PEL, Peruvian in Lima; PUR, Peurto Rican in Peurto Rico. CDX, Chinese Dai in Xishuangbanna; CHB, Han Chinese in Beijing; CHS, Southern Han Chinese; JPT, Japanese in Tokyo, KHV, Kinh in Ho Chi Minh City Vietnam. CEU, Utah residents Northern and Western European Ancestry; FIN, Finnish in Finland; GBR, British in England and Scotland; IBS, Iberian populations in Spain; TSI, Toscani in Italy. BEB, Bengali in Bangladesh; GIH, Gujarati in Houston; ITU, Indian Telugu in the UK; STU, Sri Lankan Tamil in the UK; PJL, Punjabi in Lahore Pakistan.

We used rs10488763 as a genetic proxy to test whether FDX1 impacts other human traits or diseases, including traditional risk factors for CVD. Although 3BH5C plasma levels correlated inversely with lower BMI, BP, triglyceride, fasting glucose, and HbA1c (Bonferroni-corrected P<0.05, **Supplementary Fig. 5**), we found no evidence for association of rs10488763 with these traits (P>1×10^-4^) in Biobank Japan (**Supplementary Fig. 6**; https://pheweb.jp/), UK Biobank, FinnGen, or other studies reported in the GWAS Catalog (https://genetics.opentargets.org/). rs10488763 is also not associated with non-cardiovascular diseases in these datasets. There was also little evidence for association of rs10488763 with nuclear magnetic resonance (NMR) spectroscopy-based serum metabolomic profile in 12,120 Asian and 456,571 European participants from the UK Biobank study^26^ (**Supplementary Table 10**). Genetic variants in FDX1 thus appear to have a specific effect on cardiovascular disease, that is independent of, and not mediated by, traditional risk factors. Furthermore, there is no evidence for pleiotropic effects on non-cardiovascular traits. These observations provide support for the potential safety of FDX1 as a therapeutic target for CVD risk reduction.

We observe strong global variation in allele frequencies of these genetic variants across populations (**Fig. 3c**) – allele T of rs10488763, that is associated with reduced 3BH5C and increased risk of CAD, is more common in Asian populations compared to European and African populations (minor allele frequencies in 1000G: East Asians = 0.37, South Asians = 0.25, Europeans = 0.11, Africans = 0.04), consistent with our observation of a more important role for FDX1 in the aetiology of cardiovascular disease in Asian people.

### Colocalization with gene expression

An examination of rs10488763 and rs1443120 in eQTL (expression quantitative trait loci) databases, such as GTEx and eQTLGen, indicated that these genetic variants were cis-eQTLs for *FDX1*, *RDX*, and *ZC3H12C*. To follow-up on this, we applied the summary-based MR (SMR) method^27^ to screen for *cis*-gene expression signatures (GTEx v8, N=670) associated with plasma 3BH5C levels. We found significant association between 3BH5C levels and expression of *cis-*gene *FDX1* (top variant rs2846734, P_SMR_ = 8.5×10^-10^, P_HEIDI_ = 0.23; **Supplementary Table 11**).

Association with *RDX* while significant, failed the HEIDI (heterogeneity in dependent instruments) test (P_SMR_ = 6.8×10^-10^, P_HEIDI_ = 0.008), and association with *ZC3H12C* was not significant (P_SMR_ > 0.05). To evaluate if the identified pair of traits (3BH5C-*FDX1*) share a single causal variant, we performed pairwise colocalization analysis (COLOC)^28^ using whole blood eQTL data from both HELIOS (N=1,228) and public datasets (GTEx v8, N=670). We found strong evidence for colocalization between 3BH5C and *cis*-gene *FDX1* under a single causal variant assumption using both HELIOS and GTEx data (PP H4 = 0.58 and 0.72, respectively; **Fig. 4**).

**Figure 4.**
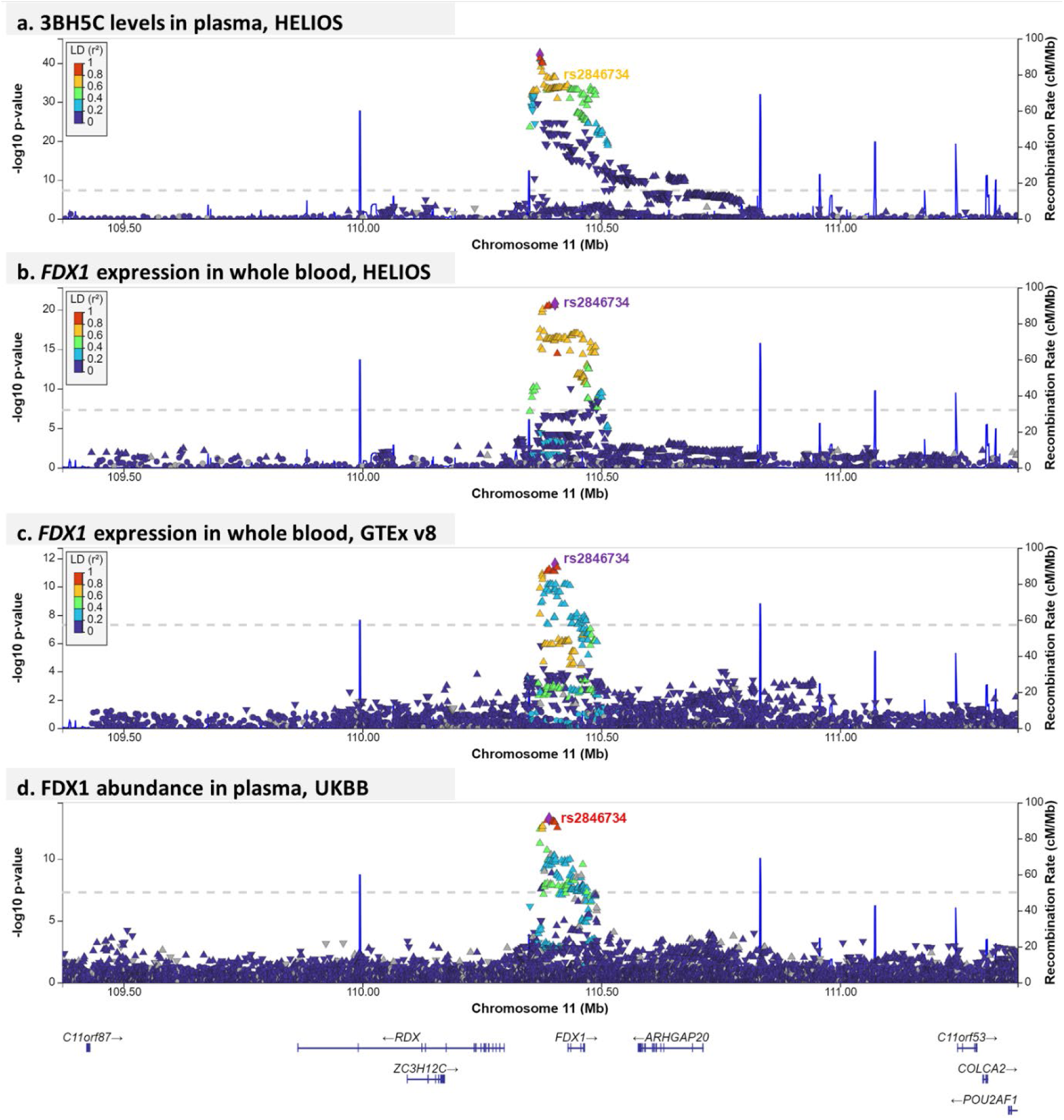
Colocalization of genetic variants associated with plasma levels of 3BH5C and expression of *FDX1*. (a) Chromosome 11 regional plot showing genetic variants associated with plasma levels of metabolite 3BH5C in HELIOS. (b) Chromosome 11 regional plot showing genetic variants associated with expression of *cis*-gene *FDX1* in whole blood in HELIOS (colocalization posterior probability=0.58). (c) Chromosome 11 regional plot showing genetic variants associated with expression of *cis*-gene *FDX1* in GTEx v8 Whole Blood tissue (colocalization posterior probability= 0.72). (d) Chromosome 11 regional plot showing genetic variants associated with plasma protein abundance of FDX1 in UKBB (colocalization posterior probability= 0.87). In all four regional plots, each dot represents a genetic variant, the genomic positions based on GRCh38 are on the x-axes and −log_10_ P-values of associations are on the y-axes. Genes annotated to this region are shown at the bottom of the panel. The top *cis*-eQTL of *FDX1*, rs2846734 is highlighted. The colour of variants indicate their linkage disequilibrium (LD r^2^) with the top variant in each panel. 1000 Genomes East Asian population served as the LD reference panel for HELIOS panels, 1000 Genomes European population served as LD reference panel for GTEx panel, and the overall 1000 Genomes population served as LD reference panel for UKBB panel which utilized the combined *cis*-pqTL summary data. Figures were generated using LocusZoom. 3BH5C, 3beta-hydroxy-5-cholestenoate.

To build on this work, we also tested for colocalization of genetic variants associated with protein expression of *FDX1* in plasma and 3BH5C levels. We performed pairwise colocalization analysis (COLOC) using plasma *cis*-pQTL summary data from the UKBB Pharma Proteomics Project^29^ (N=49,748) and confirmed evidence of a shared causal variant (PP H4 = 0.87).

### Functional evaluation of FDX1

3BH5C is synthesized from 27-hydroxycholesterol (27-HC), through the enzymatic activity of sterol 27-hydroxylase (encoded by *CYP27A1*) in the acidic pathway of bile acids biosynthesis. Ferredoxin-1, encoded by *FDX1*, is reported to play an important role in regulating 27-hydroxylase activity by facilitating the transfer of electrons from NADPH to CYP27A1^30^. Given that *CYP27A1* is highly expressed in both hepatocytes and macrophages (https://www.proteinatlas.org/ENSG00000137714-FDX1), we next examined if FDX1 activity is required for 3BH5C synthesis in these cells. Employing CRISPR/Cas9, we specifically knocked out *FDX1* in three cell lines: human hepatoma Huh7 cells, mouse hepatocyte-derived AML12 cells, and human monocyte-derived THP1 cells (**Fig. 5a**). Notably, depletion of FDX1 in hepatocytes led to a 82-90% reduction of secreted 3BH5C levels, while cellular 3BH5C was undetectable (**Fig. 5b-5c**). In THP1-derived macrophages, we also found a substantial 72% reduction of 3BH5C levels in the knockout cells compared to wild-type control cells (**Fig. 5d**). Importantly, while 3BH5C level in wild-type macrophages was significantly elevated upon treatment with excessive cholesterol, it remained unchanged in the cells lacking *FDX1* (**Fig. 5d**).

**Figure 5.**
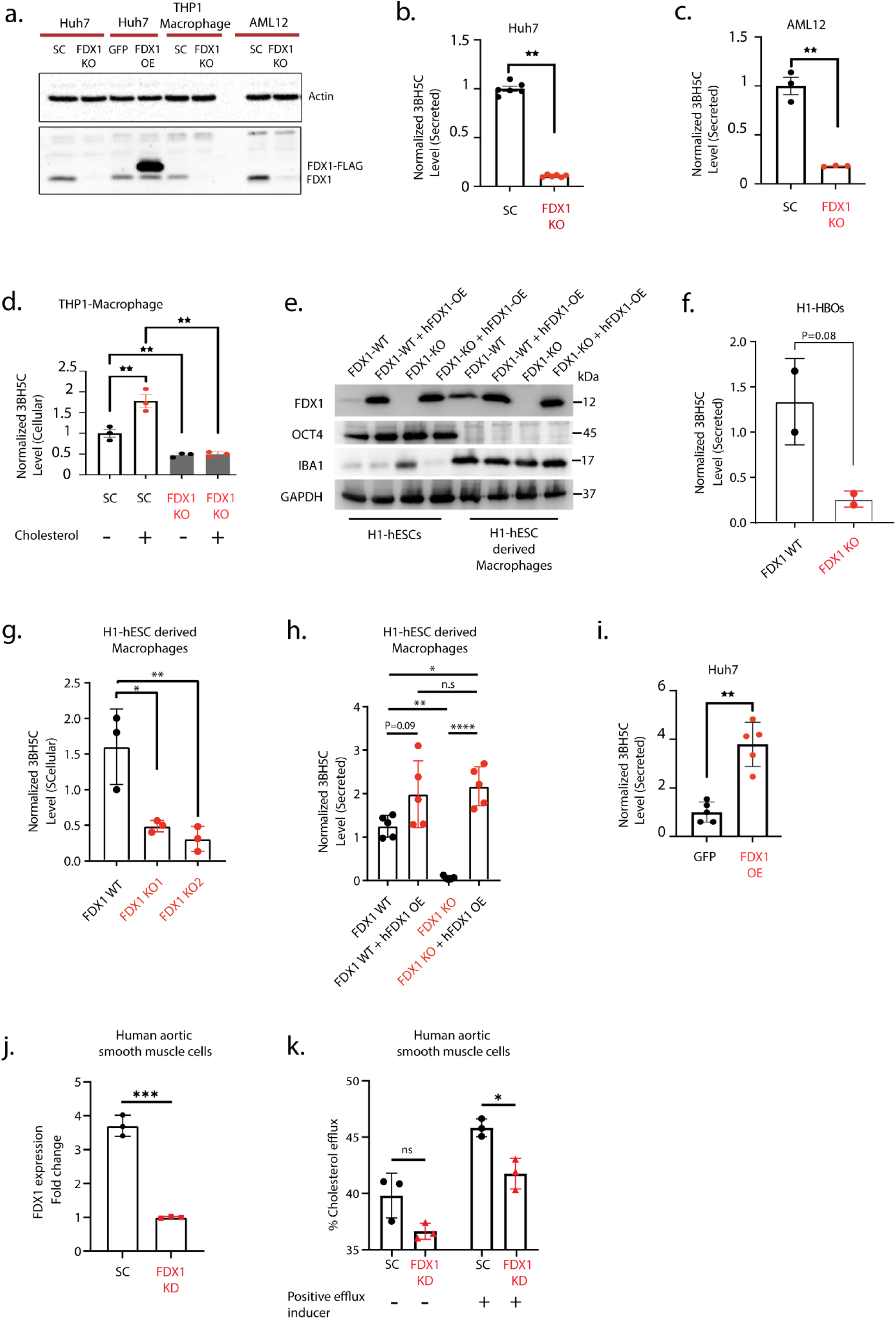
Functional validation of FDX1 as a key regulator of 3BH5C synthesis. (a) FDX1 protein expression in *FDX1* knock-out (*FDX1* KO) and *FDX1* over-expression (*FDX1* OE) human hepatoma Huh7 cells, *FDX1* KO mouse hepatocyte-derived AML12 cells, and *FDX1* KO human monocyte-derived THP1 cells, compared to scrambled controls (SC). Metabolite 3BH5C levels in *FDX1* KO cells compared to SC in (b) human hepatoma Huh7 cells, (c) mouse hepatocyte-derived AML12 cells, and (d) human monocyte-derived THP1 cells with and without cholesterol treatment. (e) FDX1 protein expression in *FDX1* WT and KO, followed by over-expression (OE) and rescue, show robust and stable transgene expression following macrophage differentiation. Measurement of FDX1 levels, and metabolite 3BH5C levels in *FDX1* KO compared to wild-type (WT) in (f) H1-derived hepatobiliary organoids (H1-HBOs), and (g) H1-derived macrophages. (h) Measurement of 3BH5C levels upon *FDX1* OE and recovery in H1-derived macrophages compared to wild-type (WT). (i) Measurement of 3BH5C levels upon *FDX1* OE in Huh 7 cells compared to GFP controls. (j) Quantitative RT-PCR of *FDX1* expression in *FDX1* knock-down (KD) aortic smooth muscle cells compared to scrambled control. (k) Percentage cholesterol efflux in *FDX1* KD cells compared to scrambled control with and without treatment with a positive efflux inducer. *P<0.05, **P<0.01, ***P<0.001 n.s : non significant. 3BH5C, 3beta-hydroxy-5-cholestenoate; FDX1, Ferredoxin-1

Considering the possibility that the immortalised cells might not faithfully reflect physiological conditions, we performed independent differentiation of human embryonic stem cells into the macrophage lineage, and hepatobiliary organoids, prior to 3BH5C quantification using mass spectrometry. We generated isogenic *FDX1* knockouts using two independent sgRNAs in H1-hESCs Pansio-1 Safe-harbour line for stable transgene expression as previously described^31^ (**Supplementary Fig. 7a**). We found that depletion of *FDX1* resulted in significantly reduced 3BH5C levels in both differentiated hepatobiliary organoids (HBOs) and macrophages compared to wild-type control cells (**Fig. 5f-5g**).

As our epidemiological findings indicated that increased plasma 3BH5C levels associate with increased FDX1 expression, we next interrogated if rescue or recombinant over-expression of *FDX1* would enhance secreted 3BH5C levels. Using co-transfection of BxbI and *hFDX1*-IRES-Puro donor, the *hFDX1* transgene was knocked into the Safe-harbour locus in both the *FDX1* WT and KO H1-hESCs Pansio-1 Safe-harbour line (**Fig. 5e, Supplementary Fig. 7b**). We found that over-expression of *FDX1* in the wild-type cells as well as rescued expression of *FDX1* in the knockout cells resulted in elevated 3BH5C levels that were significantly higher than the control cell lines (**Fig. 5h**). Consistently, recombinant over-expression of *FDX1* in Huh7 cells significantly increased secreted 3BH5C levels compared to GFP controls (**Fig. 5i**). These observations demonstrate that FDX1 positively regulates 3BH5C synthesis in both hepatocytes and macrophages.

Finally, we investigated the role of *FDX1* depletion on cholesterol efflux^32^. We transfected Human aortic smooth muscle cells (HAoSMCs) with either scrambled siRNA control or *FDX1* siRNA (**Fig. 5j**). The modified cells were then incubated with fluorescently labelled cholesterol followed by treatment with a positive efflux inducer treatment containing a cholesterol acceptor. We found that there was significant attenuation (9%) in cholesterol efflux in the *FDX1* depleted HAoSMCs treated with the cholesterol acceptor (**Fig. 5k**).

## Discussion

We investigated the metabolic profile of atherosclerosis in 8124 Asian individuals taking part in the HELIOS population cohort study. We found that lower plasma concentrations of 3BH5C, a cholesterol metabolite, are associated with increased cIMT independent of traditional vascular risk factors, and provide evidence for a causal relationship of this metabolic pathway to CVD. We showed that genetic variants influencing 3BH5C levels are associated with CVD, including myocardial infarction, angina pectoris, and coronary atherosclerosis in large population studies such as Biobank Japan, UK Biobank, and FinnGen (https://pheweb.jp/, https://genetics.opentargets.org/). We note that risk allele frequencies are stratified across ancestries – being up to 9 times more common in Asians and Latin Americans, than in European and African populations. The association of genetically determined changes in 3BH5C with CVD is also ∼5-6-fold stronger in Asian than European populations, indicating that 3BH5C metabolic pathways make a more important contribution to cardiovascular health in Asian populations.

The genetic variants potentially causal for influencing 3BH5C concentrations are also eQTLs for genes *FDX1*, *RDX*, and *ZC3H12C*. FDX1 is an iron-sulphur protein essential for the reduction of mitochondrial CYP450, involved in sterol, bile acid, and vitamin D synthesis^33^. RDX (radixin) is a cytoskeletal protein crucial in binding actin filaments to the plasma membrane and studies have reported a potential role of RDX in endothelial barrier function^34^. ZC3H12C is a zinc-finger protein involved in inflammatory signaling and is reported to be involved in the inhibition of endothelial activation and vascular inflammation^35^. To understand which of these genes is most likely involved with 3BH5C, we performed colocalization analysis using the SMR method, and observed that variants associated with 3BH5C are also strongly associated with *FDX1* and *RDX* but not with other proximal genes. The observed colocalization could indicate (i) one or more shared causal variants (pleiotropy or causality), (ii) two independent causal variants in strong linkage. We employed the HEIDI test in the SMR tool to distinguish between pleiotropy or causality, and linkage, and found that the causal relationship with *FDX1* expression is robust. Furthermore, using a Bayesian method, we found supporting evidence for colocalization between 3BH5C levels and *FDX1* gene expression as well as FDX1 protein expression in blood. The directions of effects observed in our analyses indicate that genetic variants which increase levels of circulating 3BH5C also increase expression of *FDX1* and the protein’s abundance in blood, and decrease risk of CAD. These findings identify a potentially protective role of 3BH5C in cardiovascular health. Chen et al. also reported a relationship between 3BH5C and CAD in the same direction in an aging Canadian cohort^17^. We not only independently confirm this relationship in an Asian population cohort, but also identify and validate the functional role of *FDX1* in the metabolism of cholesterol into 3BH5C.

3BH5C is an intermediate metabolite in the acidic pathway of cholesterol metabolism that results in the production of oxysterols and primary bile acids. Oxysterols are essential for cholesterol transport and lipid homeostasis, and are endogenous LXR agonists ^36^. Decreased circulating bile acids are reported to be predictors of CAD, primarily indicating dysregulation of reverse cholesterol transport ^37,38^. Bile acid sequestrants have been long prescribed for the treatment of hypercholesterolemia, and the potential to reduce risk of CAD^39^. Efflux of cholesterol from macrophages and peripheral tissue, and its transportation to the liver for metabolism into bile acids in the hepatocytes is believed to be a critical route for removal of excess cholesterol from the body and prevention of foam cell formation ^36,40^.

The acidic pathway for cholesterol metabolism is initiated by mitochondrial CYP27A1 that converts cholesterol to (25R)-26-hydroxycholesterol (27HC), and then further to 3BH5C, which is then rapidly metabolized by CYP7B1 to 7-alpha-hydroxy-3-oxo-cholestenoate (7-HOCA) and downstream into chenodeoxycholic acids. During these enzymatic conversions, CYP27A1 binds to FDX1, its redox partner. Mutations in FDX1-binding domain of CYP27A1 cause cerebrotendinous xanthomatosis (CTX)^41–43^. CTX is characterized by reduced activity of CYP27A1, leading to decreased bile acid synthesis, accumulation of cholesterol in macrophages and the CNS, and the development of tendon xanthomas, premature atherosclerosis and neurological disorders^44^.

Macrophages show high expression of CYP27A1, and produce modified sterols that cause LXR activation leading to subsequent upregulation of genes involved in cholesterol efflux, as well as reduced foam cell formation^45,46^. These observations are consistent with the hypothesis that eQTLs which decrease *FDX1* expression reduce CYP27A1 activity, leading to decreased levels of 3BH5C, dysregulated cholesterol efflux and the development of atherosclerosis.

We, therefore, investigated the effect of *FDX1* modulation on 3BH5C production in hepatocytes and macrophages, and cholesterol efflux in aortic smooth muscle cells. We found that, compared to wild type controls, depletion of *FDX1* in immortalized murine and human cell lines, human embryonic stem cell-derived macrophages, and hepatobiliary organoids, lead to a significant reduction in 3BH5C levels. Additionally, 3BH5C levels fail to recover upon treatment with excess cholesterol in *FDX1*-depleted macrophages. We also confirmed that over-expression of *FDX1* in human hepatocytes as well as wild-type (over-expression) and *FDX1* knock-out (rescue) human embryonic stem cell derived macrophages increase 3BH5C levels. These findings highlight the critical role of *FDX1* in 3BH5C synthesis. Finally, we demonstrated that *FDX1* knock-down in human aortic smooth muscle cells result in significant decrease in cholesterol efflux, further confirming the key role of FDX1 in this cholesterol efflux pathway. Our results extend recent observations in *FDX1* and *FDXR* deficient murine and human cells, which show the presence of altered lipid metabolism leading to accumulation of lipid droplets, and increased levels of cholesterol and triglycerides^32,47^.

Our molecular epidemiological investigations of a deeply-phenotyped Asian population study, combined with functional validation studies, demonstrate that *FDX1* is a key regulator of cholesterol metabolism via the acidic pathway and identify 3BH5C as a biomarker of its disruption. We show that *FDX1* mediated perturbation of this cholesterol efflux pathway is strongly associated with CVD risk in Asian populations, with effect sizes that are 5-6 fold higher than in Europeans.

The effect of FDX1 on CVD risk is independent of traditional risk factors, and does not appear to impact biological processes outside the cardiovascular system. Our results, thus, pave the way for future studies to further investigate the effect of dysregulation of this pathway in the pathogenesis of atherosclerosis, and explore the therapeutic potential of *FDX1* in the prevention of CVD.

## Methods

### Study design

Our experimental design is summarized in **Figure 1**. In brief, we performed a metabolome-wide association study of carotid intima media thickness (cIMT), a well-studied marker of cardiovascular vascular risk. We identified a panel of metabolites that associated with mean cIMT in the study population. Utilizing genome-wide association studies (GWAS) of these metabolites, cIMT, and coronary artery disease (CAD), we conducted Mendelian randomization studies to evaluate potential causal pathways linking the sequence variants and metabolites to disease. We identified a potentially causal association between a lipid metabolite, 3BH5C, and CAD. We then performed functional annotations, fine-mapping, and colocalization studies to further investigate the mechanisms involved, through which we identified *FDX1* as a critical component of cholesterol metabolism. Subsequently, we performed functional studies in murine and human hepatocytes and macrophages, as well as differentiated human embryonic stem cells to demonstrate that modulation of *FDX1* altered cholesterol metabolism into 3BH5C.

### Participant recruitment and collection of baseline data

HELIOS (https://www.healthforlife.sg/) is a population-based cohort consisting of 10,004 multi-ethnic Asian men and women aged 30-84 years, and living in Singapore (ethics approval by the Nanyang Technological University’s Institutional Review Board: 2016-11-030). Participants were recruited from the general population, through community outreach programs to ensure diversity in ethnicity and socio-economic background. Exclusion criteria were pregnancy/ breastfeeding, major illness requiring hospitalization /surgery, cancer treatment in the past year or recent participation in drug trials. Participant ethnicity was based on self-report, and comprised three primary ethnic groups: i. Chinese and other East Asian (‘Chinese’), ii. Malay and other South-East-Asian (‘Malay’) and iii. Indian and other South Asian from Indian subcontinent (‘Indian’).

All consenting participants completed a comprehensive structured assessment, according to a standardized protocol. The assessment included: i. health and lifestyle questionnaires; ii. physiological assessments, iii. imaging evaluations and iv. collection of fasting biological samples. General physiological measures including weight, height, waist and hip circumference and resting blood pressure were extracted. Waist and hip circumferences, and blood pressure, were assessed 3 times and averaged. Total cholesterol, low-density lipoprotein cholesterol (LDL-C), high-density lipoprotein cholesterol (HDL-C), triglycerides, glucose, and HbA1c levels were measured by an accredited laboratory (QuestLab, Singapore, SAC–SINGLAS ISO 15189:2012) using ADVIA 1800 chemistry system (Siemens Healthcare, Munich, Germany). Diabetes status was defined based on recommendations by the American Diabetes Association (ADA) and the World Health Organization (WHO)^48,49^. Participants were categorized as having diabetes if they had a fasting blood glucose of 7.0 mmol/l or greater, or an HbA1c measurement of 6.5% or greater, or they self-reported diagnosis of diabetes by a physician. Based on several international guidelines^50^ (National Institute for Health and Clinical Excellence, European Society of Cardiology, American Heart Association), participants were categorized as having hypertension if they had a systolic/diastolic blood pressure of 140/90mmHg or higher, or they self-reported diagnosis of hypertension by a physician. Smoking status was obtained from questionnaire-based self-reported data categorized as “never smoker”, “previous smoker”, and “current smoker”.

### cIMT estimation

Two-dimensional and three-dimensional carotid ultrasound imaging of the common carotid artery (CCA) was performed using a Philips iU 22 ultrasound system. As per recommendations by the American Society of Echocardiography (ASE)^51^, cIMT measurement was centred at 1 cm proximal to the carotid bifurcation (avoiding plaques), and images obtained at the lateral and posterior angles bilaterally. Images were analyzed using the Philips Qlab software for automated IMT measurements. ECG gating was used to time the measurement at the end-diastolic phase of the cardiac cycle. Manual quality check of automated measurements was performed to ensure that at least 95% of the tracing was correctly identified for the intima-media interface. Inter-operator and inter-reader variability was determined periodically to ensure that the coefficient of variation was lower than 5%. For this study, we used mean cIMT, i.e. average of all 4 measurements. Mean cIMT values were log-transformed prior to further analyses.

### Metabolite quantification

Samples collected from study participants were sent to Metabolon, Inc. (Durham, NC, USA) for untargeted metabolomic profiling on the Metabolon Discovery HD4 platform using published methods^52^. Samples were diluted with methanol, shaken and centrifuged to precipitate protein, liberate small molecules associated with or trapped in proteins, and to maximize the diversity of metabolites recovered. The resulting extracts were divided into two for analyses by two separate reverse phase (RP)/ultra-performance liquid chromatography (UPLC)-mass spectrometry (MS)/MS methods with positive ion mode electrospray ionization (ESI), one for analysis by RP/UPLC-MS/MS with negative mode ESI, one for analysis by hydrophobic interaction chromatography/UPLC-MS/MS with negative ion mode ESI, and 4 plates reserved for potential re-analysis if needed. The plates were dried under warm N2 and stored. Immediately prior to LC-MS analysis the individual plates were reconstituted in solvents specific to the designated analytical method. All analyses were performed using Waters Acquity UPLC systems plumbed to allow alternating injections on 2 columns coupled to Thermo Q-Exactive mass spectrometers. Metabolon identified compounds by using proprietary in-house software to compare experimental data to a library of authentic standards containing accurate mass, retention time, and fragmentation data as previously described^52^. A series of QC samples including technical replicates and process blanks were interspersed with the experimental samples. Process blanks consisted of aliquots of DI H2O taken through the entire analytical process while the technical replicates consisted of aliquots of pooled human plasma. These samples, along with the inclusion of both instrument performance standards and process assessment standards in each sample, were used to confirm analytical performance. The process blanks were further used to remove artifacts from the final data set (artifacts, in this case, being defined as any compound with a signal intensity <3× the signal in the process blanks).

1,696 metabolites were initially identified and the unadjusted peak area-under-curve values for each metabolite were then normalized across shipment batches using median-scaling. Metabolites with missingness >20% were excluded, and the remaining imputed with the minimum value for each metabolite under the assumption of missingness due to low abundance. There were 235 participants in the cohort for whom samples had been collected from two visits 335 days apart on an average.

Data from samples corresponding to the second visit of these participants were excluded from this study. These samples were separately utilized to evaluate the longitudinal stability of the 883 metabolites using a paired two-sided t-test. 3BH5C concentrations were not significantly different across two visits (Bonferroni-corrected P<0.05). Principal Component Analysis (PCA) was utilized to identify and exclude outliers, defined as samples that were >6 standard deviations away from the mean of the first two principal components. Finally, metabolite levels were log-transformed and scaled to a mean of 0 and standard deviation of 1 prior to further analyses. Analyzed metabolites include partially-characterized molecules, X-compounds whose identity are unknown, and metabolites that were not confirmed against a known chemical standard (denoted by an asterisk in the name).

### Metabolite association

Complete carotid imaging and metabolite data were available for 8,124 samples, with self-reported Chinese, Malay, and Indian ethnicity. Multivariate regression analysis was used to evaluate the association of metabolites with mean cIMT adjusted for age, sex, ethnicity, and batch (Model 1, N=8124). For metabolites that showed significant association with cIMT in Model 1 (P<0.05 after Bonferroni correction), we performed a multivariate analysis that included BMI, systolic BP, total cholesterol, T2D status and smoking status, as measured traditional cardiovascular risk factors, along with age, sex, ethnicity, and batch (Model 2, N=8056). As a final sensitivity analysis, we repeated Model2 in a subset of the cohort who were not on cholesterol-lowering drugs (Model 3, N=6689).

### Genetic data and quality control

We carried out whole genome-sequencing (WGS; 15x depth) on a subset of 2,400 participants, who were part of the SG10K National Precision Medicine Phase I study, using the Illumina HiSeqX platform. GATK best practices were followed for variant calling and quality control described by Wong et al.^53^. Briefly, we excluded variants that failed VQSR filter and had a read depth <5x. We also removed samples that had abnormal ploidy, cross contamination rate >2%, error rate >1.5%, call rate <95%, and ti/tv ratio, het/hom ratio, and in/del ratio >6 ethnicity-stratified median average deviations. We then used the TopMed Imputation Server to impute autosomal variants to the TopMed (vR2) reference panel using the EAGLE2+Minimac4 pre-phasing and imputation pipeline^54^. Genetic variants with allele frequency differences >0.20 between the TopMed reference panel, palindromic variants with allele frequency >0.4 and genotyped variants that are not in the TopMed reference panel were excluded using PLINK v1.90. The Robust Unified Test for Hardy-Weinberg equilibrium (RUTH) method^55^ was applied to the raw variant call format (VCF) files for each chromosome in all individuals in order to identify genetic variants that fail Hardy-Weinberg equilibrium (HWE) while accounting for population structure. HWE test statistics were generated using the genotype likelihoods (*--field PL* command) and the top four genotypic principal components (*--num-pc 4* command) to account for population structure, with HWE P-values calculated using the likelihood ratio test (*--lrt-em* command).

Genetic variants with RUTH HWE test P <10^-6^ were excluded. Additional pre-imputation quality control excluded autosomal genotyped variants with MAF <0.01 and variant missingness call rate >5% within the three main genetically-inferred ancestral groups (Chinese, Indians and Malays) using PLINK v1.90, with genetic variants at the intersection brought forward for imputation. A total of 3,245,000 autosomal variants survived quality control and were used as input for imputation. Approximately 285 million autosomal variants were available following imputation. Post-imputation quality control excluded imputed variants with MAF <0.001 in at least one of the three main genetically-inferred ancestral groups (Chinese, Indians and Malays), as well variants with imputation INFO score <0.30 and RUTH HWE test P <10^-3^. A total of 7,101,506 imputed autosomal variants formed the final dataset. Autosomal genetic relationship matrix (GRM) between individuals was calculated from the full set of imputed variants using the *--make-grm* command in GCTA v1.93. Unrelated individuals within each of the ancestral groups were identified with off-diagonal elements of the genetic relationship matrix of less than 0.10 (i.e., equivalent to excluding approximately third-degree relatives or closer) using the *--grm-cutoff* command in GCTA v1.93. Finally, ancestral outliers were defined as individuals more than three times the inter-quartile range (IQR) from the median of the first two genotypic principal components calculated from the GRM using the *–pca* command in GCTA v1.93.

### Genome-wide association study

Genome-wide association analyses for all metabolites was performed on the quality-controlled set of 1,876 HELIOS participants with metabolite data. Each metabolite was adjusted for age and residuals standardized to z-scores in each sex and ancestral group, removing mean and variance differences between the sexes and between ancestral groups. This quality-controlled set of metabolites were used for downstream analyses unless indicated otherwise. Further, each metabolite was modelled as a linear function of the number of reference alleles in a leave-one-chromosome out linear mixed regression framework using the --*mlma-loco* command in GCTA v1.93. The model for each metabolite can be written as,

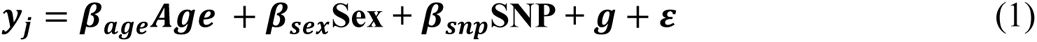

where, ***y*** is the metabolite level of the ***j***^*th*^ metabolite (following quality control; see above), with sample size *n*; ***β*** is the fixed effect estimate; ***SNP*** is a genetic variant coded as 0, 1, or 2; ***g*** is the total genetic effect for all the individuals where, 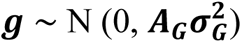, and ***A***_***G***_ is the autosomal GRM calculated from the set of all autosomal imputed genetic variants excluding those on the same chromosome as the target variant (i.e. leave-one-chromosome-out); and ***ε*** is the residual. Significance was assessed with a likelihood ratio test statistic and a P-value calculated by comparing the test statistic to a ***χ***^2^-distribution with one degree-of-freedom. To assess inflation of the test statistic due population substructure, we calculated the genomic inflation factor as the ratio of the observed versus expected median ***χ***^2^ statistic, and by applying LD Score regression (LDSC) to the metabolite GWAS summary statistics to calculate the LDSC intercept, where a LDSC intercept that deviates from unity may indicate bias due to population substructure or cryptic relatedness.

### Fine-mapping and functional annotation

R package susieR (0.12.27) was used for fine-mapping (i) 500kb (±250kb), (ii) 1mb (±500kb), (iii) 3mb (±1.5mb) regions centred around the sentinel variant to generate 95% credible sets. This fine-mapping approach (Sum of Single Effects) is based on a sparse multiple regression model that implements a modified Bayesian method for stepwise forward selection called "Iterative Bayesian Stepwise Selection" (IBSS) that computes approximate posterior distributions of variables at each step. Credible sets are generated based on these posterior distributions and have a high probability of containing causal variables with non-zero effect.

The Functional Mapping and Annotation (FUMA)^24^ web tool v1.4.1 was used for functional annotation of genetic variants and interpretation of GWAS results. Clumping thresholds used for identification of lead variants were P=5×10^-8^, r^2^≥ 0.05, and LD block window = 500kb. All other parameters were set to their default values, and reference population used was 1000G. Candidate variants defined as P<0.05 and r^2^≥ 0.05 with any of the identified lead variants were included for further annotations including ANNOVAR for functional consequence, RegulomeDB scores for evidence of regulatory activity, and 15-core chromatin state mapping across 127 tissue/cell types (RoadMap + ENCODE epigenomes).

### Phenome-wide association testing of genetic variant with NMR-based metabolites

We tested association of 168 NMR-based metabolite biomarkers with the candidate SNP, rs10488763, in the full UK Biobank within principal component-assigned ancestry groups^26^. Biomarker values were log(1 + x) transformed, and values more than 4 standard deviations from the mean were removed. Imputed genotypes (version 3) were downloaded from the UK Biobank website. We tested the association between genotype and transformed biomarker value in a linear model, with age, sex and 10 genotype principal components included as covariates. Ancestry groups were assigned by projecting UK Biobank samples onto principal components generated using genotypes from the 1000 Genomes samples (release 20130502), and using a random forest classifier (trained on 1000 Genomes continental ancestry labels) to assign ancestry group based on the first 4 principal components. Principal component analysis was carried out in PLINK v2.0, and the classifier was built using the R package randomForest. Model fitting was implemented using the function fastLm in the R package RcppEigen. This analysis was carried out under approved UK Biobank project ID 30418 by Nightingale Health.

After QC, total sample sizes used in the analysis ranged (depending on biomarker) from 424,165-456,571 for Europeans, 2,363-2,535 for East Asians, 9,195-9,585 for South Asians. We then performed a fixed-effects meta-analysis of EAS and SAS population-specific summary statistics using the R package metafor (4.6) to generate combined meta-analyzed results for “Asian” population. A P-value threshold of 0.0003 (correcting for the 168 metabolites tested) was used to assess significant associations.

### Mendelian randomization (MR)

Generalized summary-based Mendelian randomization (GSMR)^13^ was performed to identify any potential causal relationship between the 252 circulating plasma metabolites associated with mean cIMT (exposure) and risk of atherosclerosis (outcome). Specifically, two outcomes were investigated: (1) cIMT and (2) CAD risk. We selected the largest and most recent GWAS summary statistics for the outcomes : Yeung et al. 2022 for mean cIMT in 45,185 individuals in UK Biobank (UKBB)^14^ and Koyama et al. 2020 for CAD in 168,228 individuals in Biobank Japan (BBJ)^15^. The GSMR tool^13^ in GCTA v1.93 was utilized to perform MR, selecting genetic instruments of exposure at P<1×10^-5^, clumping window=1mb, r^2^=0.05, and LD reference population=1000G. Horizontal pleiotropic variants were removed using the HEIDI outlier filter at the default P<0.01. Genetic variants having a large difference in allele frequencies (greater than 0.5) between the GWAS summary statistics and LD reference dataset were removed. A suggestive GWAS significance threshold (P<1×10^-5^) was used to maximize the number of genetic instruments available for the analysis (minimum genetic instruments =10). Metabolites of interest identified to have potential causal effect on the assessed outcomes (P < 4×10^-4^ accounting for Bonferroni-correction for multiple testing), were further evaluated for robustness of findings.

To this end, GWAS summary statistics for exposure (metabolites) and outcomes (CAD) from independent cohorts were meta-analyzed by sample-size weighting using METAL, and two sample MR conducted. For each metabolite of interest, we meta-analyzed GWAS summary statistics from the 2023 CLSA (N=8,299) study^17^ and the 2022 METSIM (Metabolic Syndrome in Men, N=6,136) study^18^, both of which utilized the same Metabolon DiscoveryHD4 platform as HELIOS. For CAD, we meta-analyzed GWAS summary statistics from the 2015 study in CARDIoGRAMplusC4D (N=184,305)^20^, and the 2018 study in UKBB (N=296,525)^19^.

TwoSampleMR (0.5.6) R package was utilized to identify genetic instruments (variants with association p<5×10^-8^, clumping window=1mb, r^2^=0.05, reference population=1000G EUR), harmonize genetic variants to the forward strand (inferred from MAF) and exclude ambiguous genetic variants (for which forward strand could not be inferred). *F*-statistics were calculated for each genetic instrument to evaluate weak instrument bias, and only included in analysis if *F-statistic* >10.Three methods were implemented to evaluate potential causality: inverse-variance-weighted (IVW) MR, weighted-median MR, and weighted-mode MR. In the absence of horizontal pleiotropy, we also estimated causal effect using MR-Egger. To test for horizontal pleiotropy, we performed egger-intercept test, leave-one-out MR analysis and MR-PRESSO global and outlier tests. IVW MR analysis with CAD risk as outcome and metabolite as exposure were also conducted to evaluate potential reverse causality. Heterogeneity test for IVW-based MR estimates from Asian and European populations was conducted using the R package ‘metafor’.

### Colocalization analysis

We performed summary data–based Mendelian randomization (SMR) to test for shared genetic variants that associated with altered levels of the circulating metabolite 3BH5C, as well as expression of *cis*-genes. The SMR v1.3.1 tool^27^ was implemented using 3BH5C GWAS summary statistics from HELIOS and Whole Blood cis-eQTL (expression quantitative trait loci) summary statistics from GTEx V8 for 6433 probes (reference population1000G). This method relies on a Mendelian randomization-based approach to determine association of gene expression with a complex trait owing to pleiotropy. The HEIDI (Heterogeneity in Dependent Instruments) method was applied to distinguish between horizontal pleiotropy and linkage (HEIDI p-value threshold of 0.01). For metabolite-gene associations with Bonferroni-corrected P_SMR_ < 0.05 and P_HEIDI_ < 0.01, we tested for colocalization using the R package coloc (5.1.0)^28^, which is a Bayesian method that estimates the posterior probability of a single shared genetic variant associating with both gene expression and trait of interest. We selected a 1mb window around the sentinel variant and performed pairwise colocalization analysis between 3BH5C (HELIOS GWAS) and identified *cis*-gene target (Whole Blood *cis*-eQTL GTEx v8) under the assumption of a single causal variant and using the default prior probabilities (p1=1×10^-4^, p2=1×10^-4^, p12=1×10^-5^). We also performed colocalization analysis using the same parameters with *cis*-eQTL data from HELIOS as well as *cis*-pQTL summary data (combined population) from the UKBB Pharma Proteomics Project^29^. eQTL data in the HELIOS study were generated from RNA sequence data on 1,228 plasma samples modelled using Matrix eQTL (R package MatrixEQTL) adjusting for covariates age, gender, ethnicity, RIN (RNA integrity number) and the top 6 PEER (Probabilistic Estimation of Expression Residuals) factors^56^. Regional plots were generated using LocusZoom^57^.

### Cell lines and cell culture

All experiments performed with human ESCs using the WA01 (H1) cell line were under the supervision and guidelines of National University of Singapore institutional review board (NUS-IRB) committee.

The Huh7 cell line is a hepatocyte-derived carcinoma cell line that originated from a liver tumor in a 57-year-old Japanese male (JCRB Cell Bank, JCRB0403). The AML12 (alpha mouse liver 12) cell line was established from hepatocytes obtained from a male mouse (CD1 strain, line MT42) transgenic for human TGF alpha (ATCC, ATCC® CRL-2254™). The THP-1 cell line is a monocyte line isolated from peripheral blood from an acute monocytic leukemia patient (ATCC TIB-202™). HEK293T cells were obtained from the American Type Culture Collection. Human H1 (WA01) hESC line were obtained from WiCell Research Institute. Briefly, Huh7 and HEK293T cells were cultured in Dulbecco’s Modified Eagle Medium (DMEM) supplemented with 10% fetal bovine serum (FBS) and 1% penicillin/streptomycin. AML12 cells were cultured in DMEM/Nutrient Mixture F-12 Ham with 10% FBS, 1% penicillin/streptomycin, 0.005 mg/mL insulin, 0.005 mg/mL transferrin, 5 ng/mL selenium, and 40 ng/mL dexamethasone.

THP-1 cells were cultured in RPMI-1640 medium with 10% FBS and 1% penicillin/streptomycin. For assays, THP-1 cells were cultured in full medium and supplemented with 100nM Phorbol 12-myristate 13-acetate (PMA) (Sigma-Aldrich, P8139-1MG) for 24 hours to induce differentiation into macrophages. Afterwards, differentiated macrophages were treated with or without lipid mixture (1%) containing free cholesterol (Thermo Fisher, 11905031) for 24 hours. Medium was then replaced with basal RMPI-1640 medium with 0.3% fatty acid free Bovine Serum Albumin (Sigma-Aldrich, A8806-5G). After 16 hours, cells and medium were harvested for LC/MS.

ESC H1 lines were cultured on Geltrex-coated six-well plates at 1:200 dilution and maintained in mTesR1 medium until 80% confluency before differentiation into hepatobiliary organoids and derived-macrophages. Cells were passaged using RelesR for 5 min, before resuspension in mTesR1 medium and re-plated by a factor of 1:10. All cell lines used in this study were cultured at 37 °C under 5% CO2.

### CRISPR-mediated knockout and overexpression of *FDX1* in Huh7, AML12, and THP-1 cells

The protocol for CRISPR-based knockout of *FDX1* is adapted from a previously published paper with slight modifications^58^. In brief, cell lines with stable expression of Cas9 were generated through infection of lentiCas9-Blast (Addgene, #52962) packaged in lentivirus for 16 hours, followed by selection with 30 μg/ml blasticidin (ThermoFisher, A1113903) for 5 days.

Subsequently, guide RNAs targeting *FDX1* were constructed into lentiviral vector lentiGuide-Puro (Addgene, #52563), and packaged viruses from HEK293T cells were used to infect cells expressing Cas9 for 16 h. Cells were subsequently cultured in the presence of 10 μg/ml puromycin for 5 days before downstream assays. Murine or human *FDX1* CDS was cloned into a lentiviral EF-1α driven expression vector, with a puromycin selection cassette driven by PGK promoter (Cellomics, LVR-1011). Lentivirus was produced by transfecting HEK293T cells using 10 ug lentiviral transgene plasmid, 7.5 µg of pMDLg/pRRE, 2.5 µg of pRSV-Rev, and 2.5 µg pMD2.G (Addgene #12251, #12253 & #12259), on a 10cm dish, cultured in DMEM High Glucose media (ThermoFisher Scientific #11965092), cultured in 10% FBS, supplemented with 50 µl of PEI and 3ml of Opti-MEM™ I Reduced Serum Medium (#31985070, ThermoFisher Scientific). Following the next 48 hours, medium was changed to 5% FBS culture in DMEM media, and supernatant was collected, pooled and filtered through a PES 0.45um filter, and viral particles were concentrated using Lenti-Pac™ Lentivirus Concentration Solution (LPR-LCS-01, GeneCopoeia™) according to the manufacturer’s instructions. Cell lines were pre-incubated in 8ug/ml polybrene prior to lentiviral transduction for 24 hours, and cells undergo selection by puromycin (1ug/ml) for at least one week before downstream assays.

For overexpression of *FDX1* in Huh7 cells, murine *FDX1* CDS was cloned into lentiCas9-Blast (Addgene, #52962) by removing Cas9 and replacing with murine *FDX1* CDS. The plasmid was packaged into lentivirus, and 2mL of the filtered virus supernatant was used to perform lentiviral transduction into Huh7 cells in culture media supplemented with 6ug/mL of polybrene for 16 hours. Virus was removed thereafter and replaced with fresh media. Cells underwent Blasticidin S HCI selection (10µg/mL) for 5 days before they were used to perform downstream assays. LentiGFP-Blast, an in-house plasmid generated using lentiCas9-Blast (Addgene #52962) as the backbone, was packaged into lentivirus and transduced into Huh7 cells using the same method as a control.

### CRISPR-mediated knockout and overexpression of FDX1 in H1 human embryonic stem cells, and in hepatocyte and macrophage cell lines

The H1 Safe-Harbour hESCs (Pansio-1) *FDX1* knockout line was generated using the Plasmid pMIA3 (Addgene plasmid #109399). CRISPR sgRNA included in sense and antisense oligonucleotides were annealed by heating to 95°C, before cooling to room temperature for 60 minutes. Both independent *FDX1* sgRNA sequence 5’-GCGCGGGCCCGGAGCAGGTA-3’ and 5’-GCGCCGACACGCTCAGCGAC-3’ were designed to target the splice junction of Exon 1 and Exon 2, and to induce a 16bp deletion on Exon 1 of the FDX1 gene locus. Western blot of FDX1 protein was performed to verify the loss of FDX1 protein. Briefly, the pMIA3 plasmid was digested with Esp3I restriction enzyme and the sgRNA sequence was ligated with T4 DNA ligase (M0202, NEB) following the manufacturer’s instructions. hESCs were dissociated using Accutase (STEMCELL Technologies, 07922), and ∼2.0 × 10^6^ cells were pelleted before resuspending with 100ul of P3 Nucleofection Primary Cell Kit solution (Lonza, V4XP-3024) and 10ug of pMIA3-*FDX1*-sgRNA plasmid were mixed in suspension. According to the manufacturer’s instruction, Nucleofection was done using the CM-113 program on the 4D-Nucleofector System (Lonza). Cells were seeded in Geltrex-coated plates supplemented with 2ml of CloneR™ (STEMCELL technologies, 05888) per well diluted in mTesR1 plus (1:10 dilution). After 2 days post-nucleofection, Cell colonies were dissociated into single cells with Accutase (STEMCELL Technologies, 07922) and RFP positive cells were FACS sorted into 96-well plate format as single monoclones in CloneR™. Single cell colonies were monitored and allowed to expand in mTesR1. Genomic DNA was extracted for genotyping and indels generated by CRISPR were validated with PCR and confirmed by Sanger sequencing. For overexpression and rescue of *FDX1* clones, the transposon donor construct carrying the puromycin resistant gene (pMIA10.53) modified from pMIA10.5, was co-transfected with an integrase (BxbI) expression construct (pMIA22) using Lipofectamine Stem Transfection Reagent (ThermoFisher Scientific, STEM00001) in mTesR1 supplemented with CloneR2 (Stem Cell Technologies, #100-0691) as previously described^31^. hESC colonies were allowed to recover in CloneR2 in mTesR1 for 48 hours, before changing into fresh mTesR1 supplemented with 1ug/ml Puromycin for hESC selection. Overexpression and Rescue of hFDX1 was validated by western blot in both WT and KO clones, before and after respective lineage differentiation.

### Differentiation into Macrophages

Hematopoietic progenitor cells derived from H1 Safe-Harbour (Pansio-1) hESC cells as previously described^59^. Briefly, undifferentiated hESCs were seeded into Geltrex-coated plates supplemented with 10uM Y-27632, before changing into fresh mTesR1 media. On Day 0, hESCs were cultured in the basal media comprising of RPMI1640 medium (ThermoFisher Scientific, 21870076), 2% B-27 (ThermoFisher, 17504044), 2mM GlutaMax (ThermoFisher Scientific, 35050061) and 60 ug/mL ascorbic acid (Sigma Aldrich, A92902), supplemented with CHIR99021 (Stem Cell Technologies, 100-1042). On Day 2, basal media was supplemented with 50 ng/ml VEGF (ThermoFisher Scientific, #100-20-1MG), and 10 ng/ml FGF2 (ThermoFisher Scientific, #100-18B-1MG). The next day, cells were dissociated using Accutase, and replated into fresh geltrex-coated plates. For the next four days, medium was replaced with basal medium supplemented with 50ng/ml VEGF, 10ng/ml FGF2 and 10uM SB431542 (Stem Cell Technologies, 72234). Following this, floating progenitor cells were replated in separate vessels, and macrophage differentiation was induced by 7-day exposure to 20 ng/ml macrophage colony stimulating factor (MCSF) (ThermoFisher Scientific, #PHC9501) in advanced DMEM/F12 (ThermoFisher Scientific, #12491015).

### Differentiation into Hepatobiliary organoids

H1 Safe-Harbour hESCs (Pansio-1) cells were grown on mTesR in 6 well plates till 80% confluency. Following which, cells were trypsinized with TrypLE (TrypLE™ Express Enzyme (1X), no phenol red Catalog number: 12604013) and seeded into 96 well plates (Akita Sumitomo Bakelite, PrimeSurface® 3D culture: Clear, Ultra-low Attachment Plates: 96 well, V bottom plates) with 10µM (1:1000) Rock Inhibitor (Y-27632) (Stem cell Technologies). The cells aggregated overnight and were subjected to 12 days of growth in hepatobiliary differentiation medium (1mM dbCAMP; 100ng/ml FGF10; 3uM CHIR99021; 2uM A83-01; 25ng/ml BMP7; 50ng/ml HGF; 50ng/ml EGF; 10mM NAD+), and were further kept for 21 days in hepatobiliary maintenance medium (2uM A83-01; 25ng/ml BMP7; 10uM DAPT; 100ng/ml FGF19; 1uM dexamethasone; 50ng/ml HGF; 50ng/ml EGF; 10mM NAD+). The media was changed once every two days by the end of which mature hepatobiliary organoids were generated.

### Sample Preparation for LC/MS

Approximately 0.5-1 x 10^6^ cells were treated with 120μL of ice-cold acetonitrile, vortexed for 10 minutes, then centrifuged at 12,000g for 10 minutes at 4°C. The supernatant was carefully transferred to an Ostro 96-well plate 25mg solid-phase extraction (SPE) cartridge (Part No. 186005518, Waters Pacific Pte. Ltd.) in conjunction with a positive pressure manifold 96 processor, PPM-96 (Agilent Technologies, Singapore) utilizing purified N2 gas. The flow through was collected on a 96-microwell plate before loading into the 5-6°C auto-sampler of the LC/MS system.

Cell media were subjected to similar organic extraction and SPE but were concentrated for the LC/MS analysis using a freeze dryer/vacuum concentration unit (Martin Christ Alpha 1-2 LDplus/ RVC 2-25 CDplus system, Osterode, Germany) equipped with a ULVAC GLD-202 (Miyazaki, Japan) oil rotary vacuum pump. Typically, 1.6-3.2mL of extracted media were concentrated and reconstituted into a smaller volume of 100-200mL acetonitrile.

### LC/MS assays

3beta-hydroxy-5-cholestenoic acid, CAS: 56845-87-5 was purchased from Merck Singapore Pte Ltd. The stable isotope internal standard 3beta-hydroxy-5-cholestenoic acid-D5, CAS: 1620239-03-3 was purchased from Avanti Polar Lipids, Inc. via Scientific Resources Pte Ltd (local distributor). Raptor C18 1.8mm 50 x 2.1mm column (Cat. No. 9304252, Restek Corp., PA, USA) was used maintaining a temperature of 50°C. Solvent A is the aqueous/buffered 4mM ammonium acetate and solvent B is methanol/acetonitrile 1:1. The percentage of each solvent used at each time was varied using the binary pump (Agilent Model G7120A) with the percentage of Solvent B as follows: 40% between 0 to 1.00 min, 80% at 6.70 min, 95% between 6.90 to 8.50 min and 40% between 8.51 to 9.50 min with a stop-time at 11.00 min. The sample injection volume was 15mL and the flow rate of the solvent system was 0.500 mL/min.

The electrospray ionization (ESI) liquid chromatography-mass spectrometry (LC/MS) assays were performed using 1290 Infinity II LC/6495 QqQ (Agilent Technologies, Singapore) and Multiple Reaction Monitoring (MRM) in the negative mode. The transitions of m/z 415.2 → m/z 415.2 and m/z 420.6 → m/z 420.6 were monitored for 3BH5C and 3BH5C-D5, respectively, with an average retention time of 5.1 minute for both. The collision energies for all transitions were 0 eV.

The ESI source parameters are as follows: gas temperature at 290°C, gas flow of 20L/min, nebulizer pressure at 35 psi, sheath gas temperature at 350°C, sheath gas flow of 12L/min, capillary voltage of 4000V, nozzle voltage of 500V, and chamber current of 0.20µA. The iFunnel parameters had a negative high-pressure RF of 90V and low-pressure RF of 60V. Acquisitions and quantitative analysis were performed using the Agilent MassHunter Workstation Acquisition (10.0.127) and Quantitative Analysis (10.0) software.

### Immunoblotting

For immunoblotting analyses, the cells were lysed in RIPA buffer containing protease and phosphatase inhibitors (Thermo Fisher). Protein levels were quantified using BCA protein assay kit (Thermo Fisher) and lysate containing equal amount of protein was subjected to SDS-PAGE, and further transferred to polyvinylidene fluoride (PVDF) membranes, followed by incubations with FDX1 primary (Protein Tech, 12592-1-AP) and secondary rabbit ECL HRP antibodies (Cytiva Lifesciences, NA934).

### siRNA-mediated FDX1 knockdown in Human aortic smooth muscle cells and cholesterol efflux assay

Human aortic smooth muscle cells (HAoSMCs, Lonza Bioscience, cat no. CC-2571, Lot no. 0000316661) were seeded in 6-well plates at a density of 5.2 x 105 cells/well using SMGM-2 medium (Lonza Bioscience, cat no. CC-3182). 24 hours post-seeding, cells underwent transfection with siRNA targeting human FDX1 (Thermo Fisher Scientific, cat no. 4392420, siRNA ID: s223529). A control group was treated with scrambled siRNA (Thermo Fisher Scientific, cat no. 4390847). siRNA stocks were prepared in 1x siRNA buffer (Dharmacon) to achieve a concentration of 20 µM, following manufacturer’s instructions.

For reverse transfection, siRNA was complexed with transfection reagent (Dharmafect-1, Dharmacon) in Opti-MEM Reduced Serum Media with GlutaMAX Supplement (Thermo Fisher Scientific, cat no. 51985034). This mixture was incubated for 20 minutes at room temperature before being added to the seeded HAoSMCs to achieve a final siRNA concentration of 25 nM in SMGM-2 medium.

24 hours post-transfection, the efficiency of siRNA knockdown in HAoSMCs was verified using quantitative reverse transcription PCR (qRT-PCR). Primers used included: FDX1 forward primer 5’-CTTTGGTGCATGTGAGGGAA-3’, reverse primer 5’-GCATCAGCCACTGTTTCAGG-3’; and GAPDH forward primer 5’-CCGTCAAGGCTGAGAACGG-3’, reverse primer 5’-CTCAGCGCCAGCATCGC-3’.

To assess the impact of FDX-1 knockdown on cholesterol efflux in HAoSMCs, a Cholesterol Efflux Assay Kit (Abcam, cat no. ab196985) was used. Post-knockdown and scrambled control HAoSMCs were reseeded in 96-well plates at a density of 105 cells/well. After 24 hours, cells were washed with serum-free RPMI 1640 medium (Gibco, cat no. 11875093) and incubated with a labelling reagent from the kit for 1 hour. Cells were then washed with serum-free RPMI medium before adding the equilibrium reagent from the kit, followed by incubation for 16 hours at 37°C in a 5% CO2 incubator in the dark. After overnight incubation, the equilibrium reagent was removed, and cells were washed with phenol red-free, serum-free RPMI 1640 medium (Gibco, cat no. 11835030).

For unstimulated wells, 100 μL of phenol red-free, serum-free RPMI 1640 medium was added. Treatment wells received either 20 μL of the positive efflux inducer treatment (from the kit) added to 80 μL of the same medium. After the addition of treatments, cells were incubated for another 4 hours at 37°C in the dark. Post-incubation, the supernatant was transferred to a 96-well white opaque plate (Thermo Fisher Scientific, cat no. 136101) and fluorescence intensity was measured at Ex/Em = 485/523 nm using a Synergy H1 microplate reader (BioTek). The percentage of cholesterol efflux was calculated by dividing the fluorescence intensity of the supernatant by the total fluorescence intensity of the supernatant and cell lysate, and then multiplying by 100.

## Data Availability

Data access request can be submitted to the HELIOS Data Access Committee by emailing for details.

## Acknowledgements

We are very grateful to the outstanding support of past and present members of the HELIOS study steering committee, operational study team, and administrative staff for driving the study and assisting in the data collection. We would also like to express our gratitude to the Nightingale Health Biobank Collaborative Group for providing GWAS summary statistics of NMR data from the UK Biobank Study. This work was supported by intramural funding from Nanyang Technological University, Lee Kong Chian School of Medicine, and the National Healthcare Group. J.C.C is supported by: the Singapore Ministry of Health and National Medical Research Council STaR funding scheme [NMRC/StaR/0028/2017], LCG funding [MOH-000271], Phase II National Precision Medicine Programme (Research Platform and Data Enablers) [NMRC/PRECISE/2020], the Singapore Agency for Science Technology and Research IAF-PP National Precision Medicine Program Phase 1A (A Population Level Genomic Infrastructure) [H17/01/a0/007], and the IAF-PP Asian Skin Microbiome Programme [H18/01/a0/016]. N.S. and J.C.C had full access to all the data in this study and take responsibility for the integrity of the data and the accuracy of the data analysis. Computational work for this study was partially performed on resources of the National Supercomputing Centre, Singapore (https://www.nscc.sg).

## Competing Interests

K.E.W, P.A.S, R.S and G.A.M are employees of Metabolon, and may hold stock/stock options in Metabolon. The rest of the authors have no conflicting interests.

## Author Contributions

J.C.C, P.E, E.R, J.N, E.S.L, J.L, J.B, and T.M conceived and designed the HELIOS study. R.D, T.M, collected phenotype data. K.E.W, P.A.S, G.A.M, and R.S carried out metabolite quantification. C.J.M.L, L. S. P, Y.M, M.A-J, T.T.T, V.G.K, Y.S, Y.L, Henry.Y, Haojie.Y, C.L.D, R. S. Y. F, K.Y.T, C.C designed and carried out the experimental studies. N.S, P.R.Jperformed the data analyses. D.L, M.Lam, M.Loh, H.K.N, T.M, D.T, X.W provided critical feedback on data analyses and interpretation of results. N.S, P.R.J, I.K, C.J.M.L, H.Y, L.S.P, T.M, P.A.S, J.C wrote the manuscript. All authors reviewed and contributed to the revision of the submitted manuscript.

## Data Availability

The HELIOS phenotype, metabolite, and genomic data used in this manuscript are protected and are not publicly available due to data privacy regulations. Data access request can be submitted to the HELIOS Data Access Committee by emailing for details. All summary level data have been made available in the supplementary tables. GWAS summary statistics of metabolites from the HELIOS study are currently available upon request and will be made publicly available through GWAS catalog upon publication of the manuscript.

GWAS summary statistics for van der Harst et al. 2018, Yeung et al. 2022, Chen et al. 2023 are available at GWAS catalog (https://www.ebi.ac.uk/gwas/), for Nikpay et al. 2015 at the CARDIoGRAMplusC4D Consortium website (http://www.cardiogramplusc4d.org/data-downloads/), for Koyama et al. 2020 at National Bioscience Database Center (https://biosciencedbc.jp/en; ID hum0014), and for Yin et al. 2022 at the METSIM Metabolomics PheWeb (https://pheweb.org/metsim-metab/).

## Code Availability

Genetic analysis was performed using GCTA v1.93 (https://yanglab.westlake.edu.cn/software/gcta/#Overview) and PLINK v1.90 (https://www.cog-genomics.org/plink/). FUMA annotation was performed using the webtool v1.4.1 (https://fuma.ctglab.nl/). Fine-mapping was performed using R package susieR (0.12.27) (https://cran.r-project.org/web/packages/susieR/vignettes/finemapping_summary_statistics.html). Mendelian randomization was performed using GSMR in GCTA v1.93 (https://yanglab.westlake.edu.cn/software/gcta/#MendelianRandomisation) and R package TwoSampleMR (0.5.6) (https://mrcieu.github.io/TwoSampleMR/articles/introduction.html).

Colocalization analysis was performed using SMR v1.3.1 for Linux (https://yanglab.westlake.edu.cn/software/smr/#Overview) and R package coloc (5.2.1) (https://cran.r-project.org/web/packages/coloc/vignettes/a01_intro.html). Regional plots were generated using LocusZoom (https://my.locuszoom.org/). Other analyses and plotting were performed using the following R packages: data.table (1.14.2), dplyr (1.0.9), forestplot (3.1.1), ggplot2 (3.4.2), ggrepel (0.9.1), metafor (4.6-0), ppcor (1.1), stringr (1.4.0), tibble (3.1.7), tidyr (1.2.0). R version v4.2.1 or v4.2.2 was used for all data analysis in R. Analyses and plotting scripts are available in the following <u>Github repository</u>.

## Notes

### Author Declarations

Ethics committee/Institutional Review Board of the Nanyang Technological University granted ethical approval for this work (IRB-2016-11-030)

### Summary of Updates

Methods section updated to provide more details on the phenome-wide analysis of NMR-based data.

