## Supplemental Information for "Metabolome-wide association of carotid intima media thickness identifies FDX1 as a determinant of cholesterol metabolism and cardiovascular risk in Asian populations"

### Description of tables in file “Supplementary Tables.xlsx”

**Supplementary Table 1. Cohort Characteristics.**

**Supplementary Table 2. Quality control of metabolite and research phenotype data**

**Supplementary Table 3: Descriptive statistics for distribution of 883 metabolites in the overall dataset (N=8124) and stratified by ethnicity (Chinese N=5428, Indian N=1552, Malay N=1144).**
Kruskal-Wallis (KW) and Anova tests were performed to test for differences in distribution of log-transformed metabolite abundances across the three ethnic groups.

**Supplementary Table 4: Summary statistics for association analysis of 883 metabolites with mean cIMT in the overall dataset and stratified by ethnicity.**
Overall analysis was adjusted for age, sex, ethnicity, and batch (Model 1). Ethnicity-specific analysis was adjusted for age, sex, and batch.

**Supplementary Table 5: Summary statistics for 252 metabolites significantly associated with mean cIMT.**
Model 1 was adjusted for age, sex, ethnicity, and batch (N=8124). Model 2 was adjusted for age, sex, ethnicity, batch, and traditional vascular risk factors: BMI, systolic BP, total cholesterol, T2D status, smoking status (N=8056). Model 3 was conducted in a subset of the cohort not on cholesterol-lowering drugs and adjusted for age, sex, ethnicity, batch, and traditional risk factors: BMI, systolic BP, total cholesterol, T2D status, smoking status (N=6689).

**Supplementary Table 6: Summary statistics from GSMR of 126 metabolites (exposures, HELIOS study) against CAD (outcome; Koyama et al. 2020).**

**Supplementary Table 7: Summary statistics from GSMR of 126 metabolites (exposures, HELIOS study) against mean cIMT (outcome; Yeung et al. 2022).**

**Supplementary Table 8: Summary of results from TwoSampleMR.**

**Supplementary Table 9. Functional annotation of 1,259 candidate variants in the risk loci.**
RegulomeDB score is a categorical score ranging from 1a to 7, indicating the following: 1a, eQTL + transcription factor (TF) binding + matched TF motif + matched DNase footprint + DNase peak; 1b, eQTL + TF binding + any motif + DNase footprint + DNase peak; 1c, eQTL + TF binding + matched TF motif + DNase peak; 1d, eQTL + TF binding + any motif + DNase peak; 1e, eQTL + TF binding + matched TF motif; 1f, eQTL + TF binding/DNase peak; 2a, TF binding + matched TF motif + matched DNase footprint + DNase peak; 2b, TF binding + any motif + DNase footprint + DNase peak; 2c, TF binding + matched TF motif + DNase peak; 3a, TF binding + any motif + DNase peak; 3b, TF binding + matched TF motif; 4, TF binding + DNase peak; 5, TF binding or DNase peak; 6, other; 7, not available.

**Supplementary Table 10: Summary statistic of rs10488763 PheWAS against 168 NMR-based metabolites in Europeans and Asians.**

**Supplementary Table 11: Summary statistics from SMR analysis of metabolite 3BH5C GWAS in HELIOS and Whole Blood cis-eQTLs from GTEx V8 (results from chromosome 11)**

**Supplementary Table 12: Correlation of metabolite 3BH5C with other metabolites adjusted age, sex, ethnicity (results for correlations with Bonferroni-corrected P-value < 0.05 & absolute coefficient values greater than 0.2).**

### Supplementary Figures

**Supplementary Figure 1. Distribution of mean cIMT in the study population (N=8,124).**

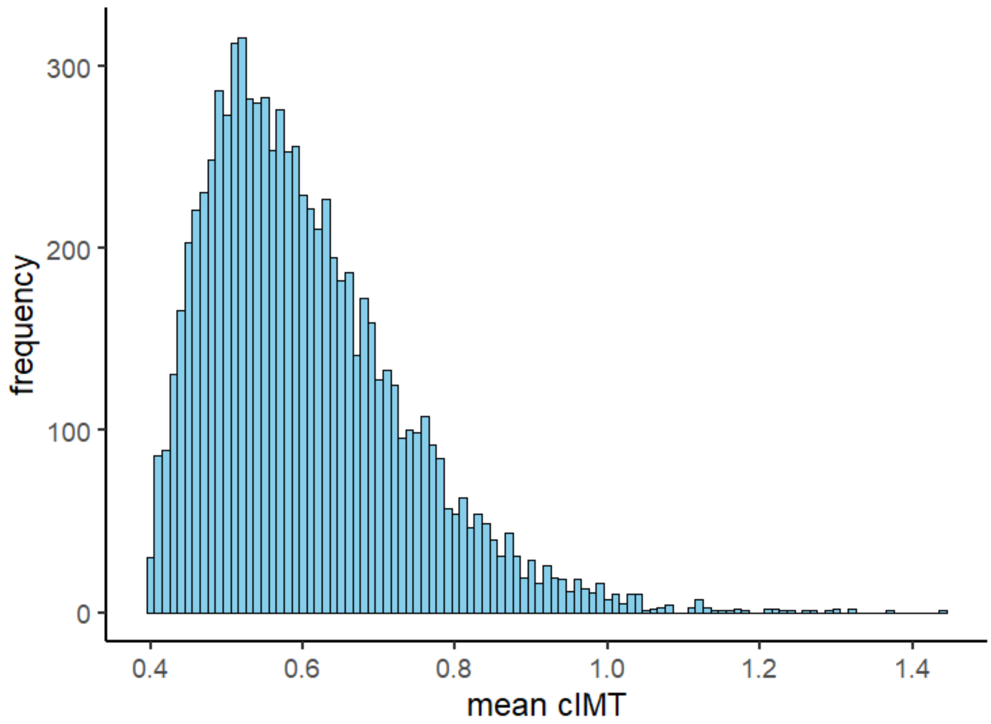

**Supplementary Figure 2. Distribution of quantified metabolites across different metabolite categories.**

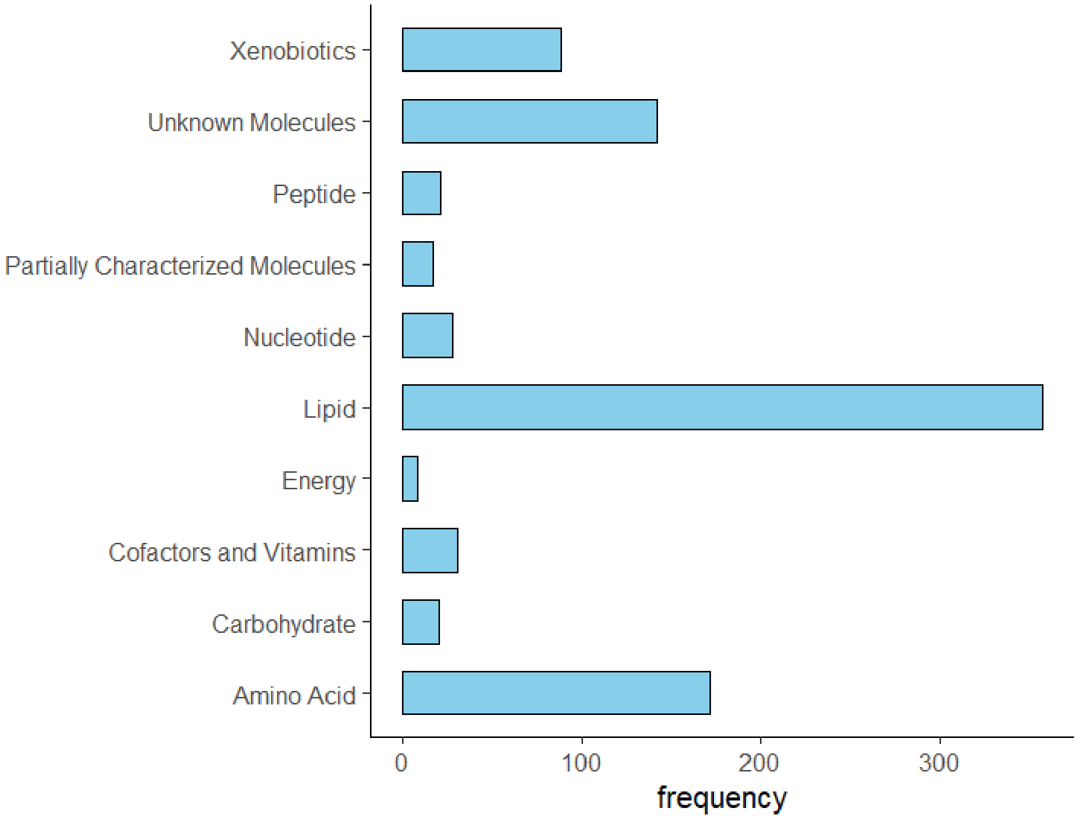

**Supplementary Figure 3. MR estimates from inverse variance weighted leave-one-out sensitivity analysis of 3BH5C (exposure) on CAD (outcome).**

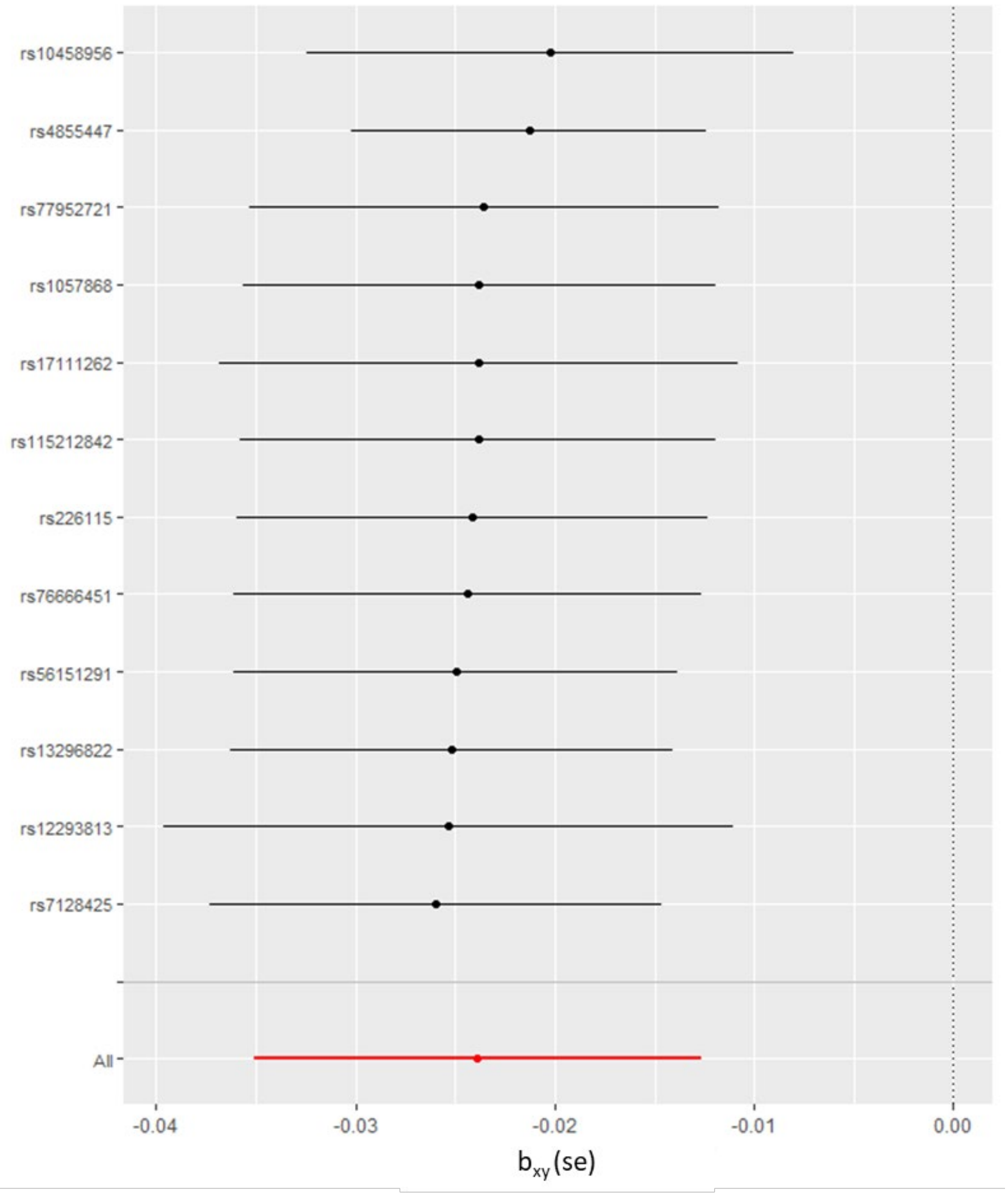

**Supplementary Figure 4. (a) Manhattan plot and (b) QQ plot of GWAS of 3BH5C in the HELIOS cohort (N=1,876; $\lambda_{GC}$=1.002).**

**a.**

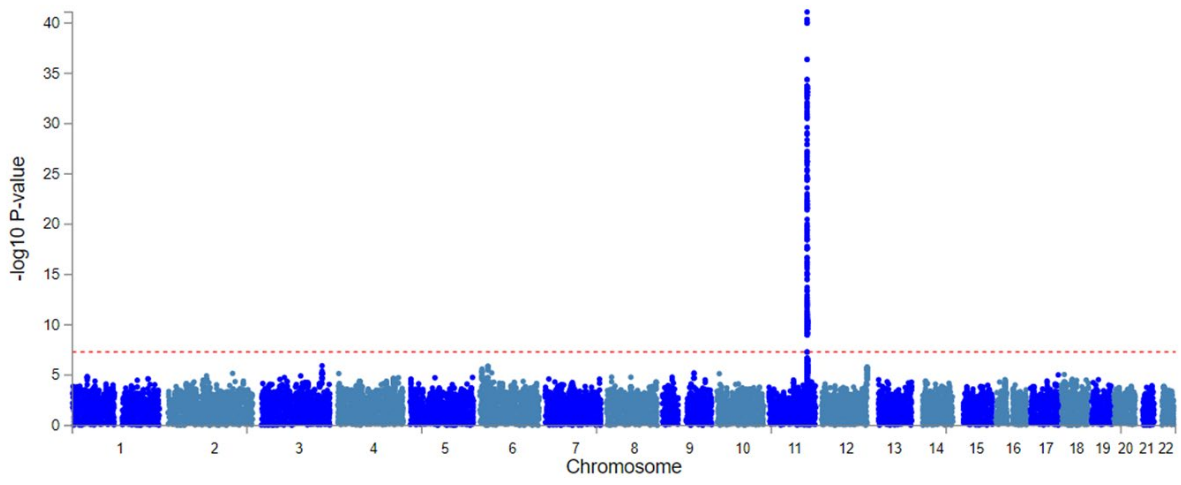


**b.**

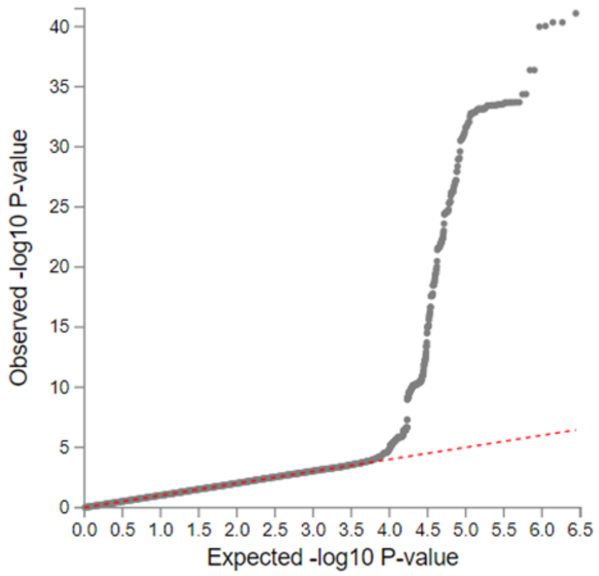


In panel (a), Manhattan plot x-axis is displaying position of genetic variants on chromosome (hg38), and y-axis is displaying the strength of association of genetic variants with mean-cIMT. Horizontal red line indicates a GWAS threshold of P=5x$10^{-8}$. The top variant is rs2051466 (P = 7.9x$10^{-43}$).

**Figure 5. Bar plots showing correlation coefficient estimates of metabolite 3BH5C with (a) traditional vascular risk factors (Bonferroni-corrected P-value < 0.05). and (b) other metabolites (Bonferroni-corrected P-value < 0.05 and absolute correlation coefficients > 0.2), adjusted for age, sex, and ethnicity.**

**a.**

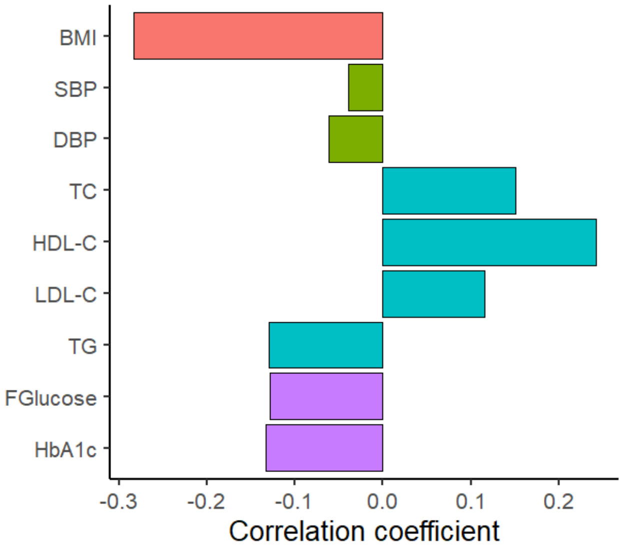

**b.**

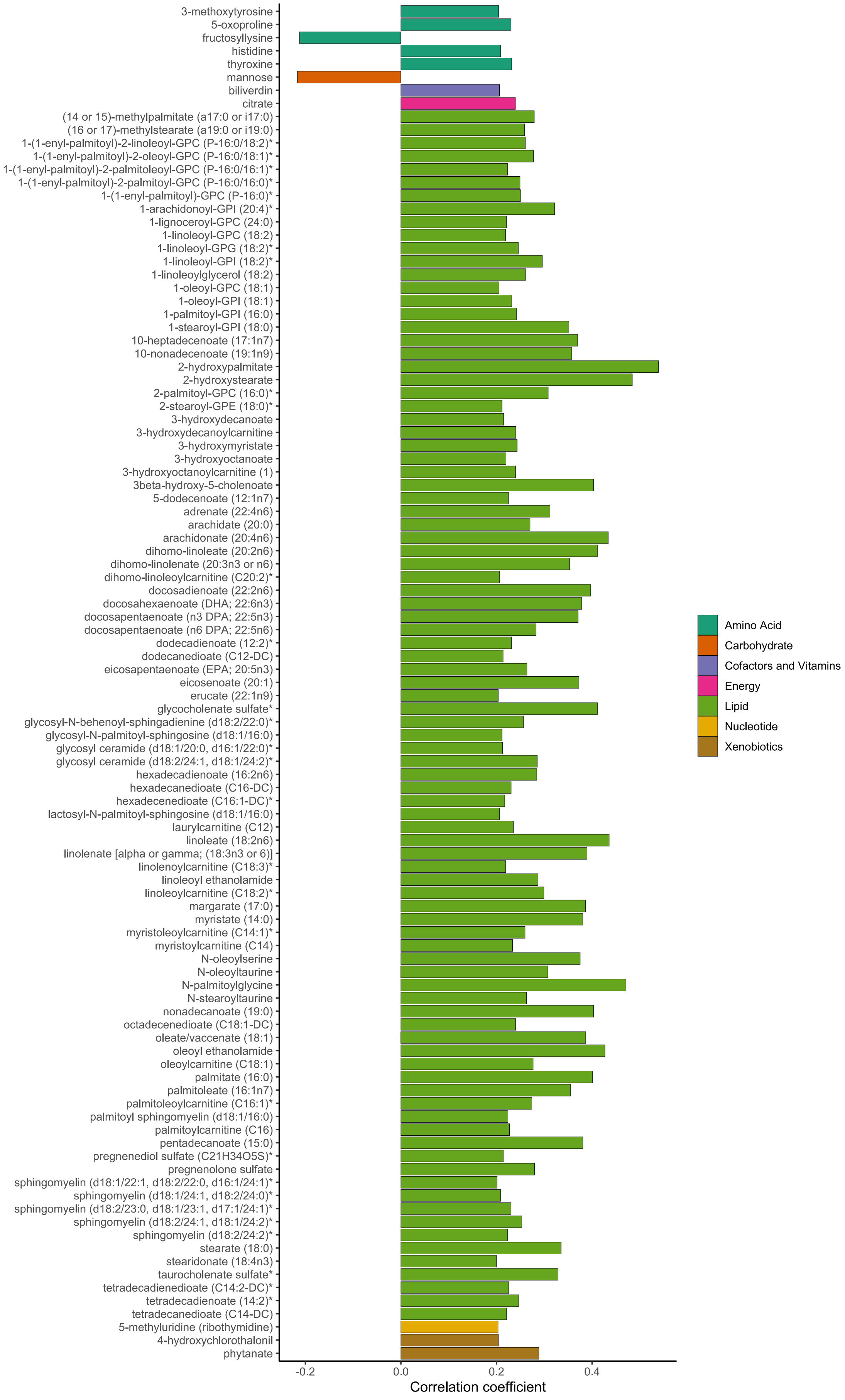
3-methoxytyrosine
5-oxoproline
fructosyllysine
histidine
thyroxine
mannose
biliverdin
citrate
(14 or 15)-methylpalmitate (a17:0 or i17:0)
(16 or 17)-methylstearate (a19:0 or i19:0)
1-(1-enyl-palmitoyl)-2-linoleoyl-GPC (P-16:0/18:2)*
1-(1-enyl-palmitoyl)-2-oleoyl-GPC (P-16:0/18:1)*
1-(1-enyl-palmitoyl)-2-palmitoleoyl-GPC (P-16:0/16:1)*
1-(1-enyl-palmitoyl)-2-palmitoyl-GPC (P-16:0/16:0)*
1-(1-enyl-palmitoyl)-GPC (P-16:0)*
1-arachidonoyl-GPI (20:4)*
1-lignoceroyl-GPC (24:0)
1-linoleoyl-GPC (18:2)
1-linoleoyl-GPG (18:2)*
1-linoleoyl-GPI (18:2)*
1-linoleoylglycerol (18:2)
1-oleoyl-GPC (18:1)
1-oleoyl-GPI (18:1)
1-palmitoyl-GPI (16:0)
1-stearoyl-GPI (18:0)
10-heptadecenoate (17:1n7)
10-nonadecenoate (19:1n9)
2-hydroxypalmitate
2-hydroxystearate
2-palmitoyl-GPC (16:0)*
2-stearoyl-GPE (18:0)*
3-hydroxydecanoate
3-hydroxydecanoylcarnitine
3-hydroxymyristate
3-hydroxyoctanoate
3-hydroxyoctanoylcarnitine (1)
3beta-hydroxy-5-cholenoate
5-dodecenoate (12:1n7)
adrenate (22:4n6)
arachidate (20:0)
arachidonate (20:4n6)
dihomo-linoleate (20:2n6)
dihomo-linolenate (20:3n3 or n6)
dihomo-linoleoylcarnitine (C20:2)*
docosadienoate (22:2n6)
docosahexaenoate (DHA; 22:6n3)
docosapentaenoate (n3 DPA; 22:5n3)
docosapentaenoate (n6 DPA; 22:5n6)
dodecadienoate (12:2)*
dodecanedioate (C12-DC)
eicosapentaenoate (EPA; 20:5n3)
eicosenoate (20:1)
erucate (22:1n9)
glycocholenate sulfate*
glycosyl-N-behenoyl-sphingadienine (d18:2/22:0)*
glycosyl-N-palmitoyl-sphingosine (d18:1/16:0)
glycosyl ceramide (d18:1/20:0, d16:1/22:0)*
glycosyl ceramide (d18:2/24:1, d18:1/24:2)*
hexadecadienoate (16:2n6)
hexadecanedioate (C16-DC)
hexadecenedioate (C16:1-DC)*
lactosyl-N-palmitoyl-sphingosine (d18:1/16:0)
laurylcarnitine (C12)
linoleate (18:2n6)
linolenate [alpha or gamma; (18:3n3 or 6)]
linolenoylcarnitine (C18:3)*
linoleoyl ethanolamide
linoleoylcarnitine (C18:2)*
margarate (17:0)
myristate (14:0)
myristoleoylcarnitine (C14:1)*
myristoylcarnitine (C14)
N-oleoylserine
N-oleoyltaurine
N-palmitoylglycine
N-stearoyltaurine
nonadecanoate (19:0)
octadecenedioate (C18:1-DC)
oleate/vaccenate (18:1)
oleoyl ethanolamide
oleoylcarnitine (C18:1)
palmitate (16:0)
palmitoleate (16:1n7)
palmitoleoylcarnitine (C16:1)*
palmitoyl sphingomyelin (d18:1/16:0)
palmitoylcarnitine (C16)
pentadecanoate (15:0)
pregnenediol sulfate (C21H34O5S)*
pregnenolone sulfate
sphingomyelin (d18:1/22:1, d18:2/22:0, d16:1/24:1)*
sphingomyelin (d18:1/24:1, d18:2/24:0)*
sphingomyelin (d18:2/23:0, d18:1/23:1, d17:1/24:1)*
sphingomyelin (d18:2/24:1, d18:1/24:2)*
sphingomyelin (d18:2/24:2)*
stearate (18:0)
stearidonate (18:4n3)
taurocholenate sulfate*
tetradecadienedioate (C14:2-DC)*
tetradecadienoate (14:2)*
tetradecanedioate (C14-DC)
5-methyluridine (ribothymidine)
4-hydroxychlorothalonil
phytanate
-0.2
0.0
0.2
0.4
Correlation coefficient
Amino Acid
Carbohydrate
Cofactors and Vitamins
Energy
Lipid
Nucleotide
Xenobiotics

In panel (a), bars are coloured by risk factors that include(1) BMI: Body Mass Index [red], (2) DBP: Diastolic Blood Pressure [green], (3) SBP: Systolic Blood Pressure [green], (4) TC: Total Cholesterol [blue], (5) HDL-C: High Density Lipoprotein Cholesterol [blue], (6)LDL-C: Low Density Lipoprotein Cholesterol [blue], (7) TG: Triglyceride[blue], (8) HbA1C: Glycated hemoglobin [purple], (5) FGlucose: fasting plasma glucose [purple]. For all correlations, Bonferroni-corrected P-value < 0.05.

In panel (b), bars are coloured by the metabolite category indicated in the figure legend. This panel shows metabolites with Bonferroni-corrected P-value < 0.05 & absolute coefficient values greater than 0.2.

**Figure 6. Volcano plot showing phenome-wide association of rs10488763 in the Biobank Japan Project.**

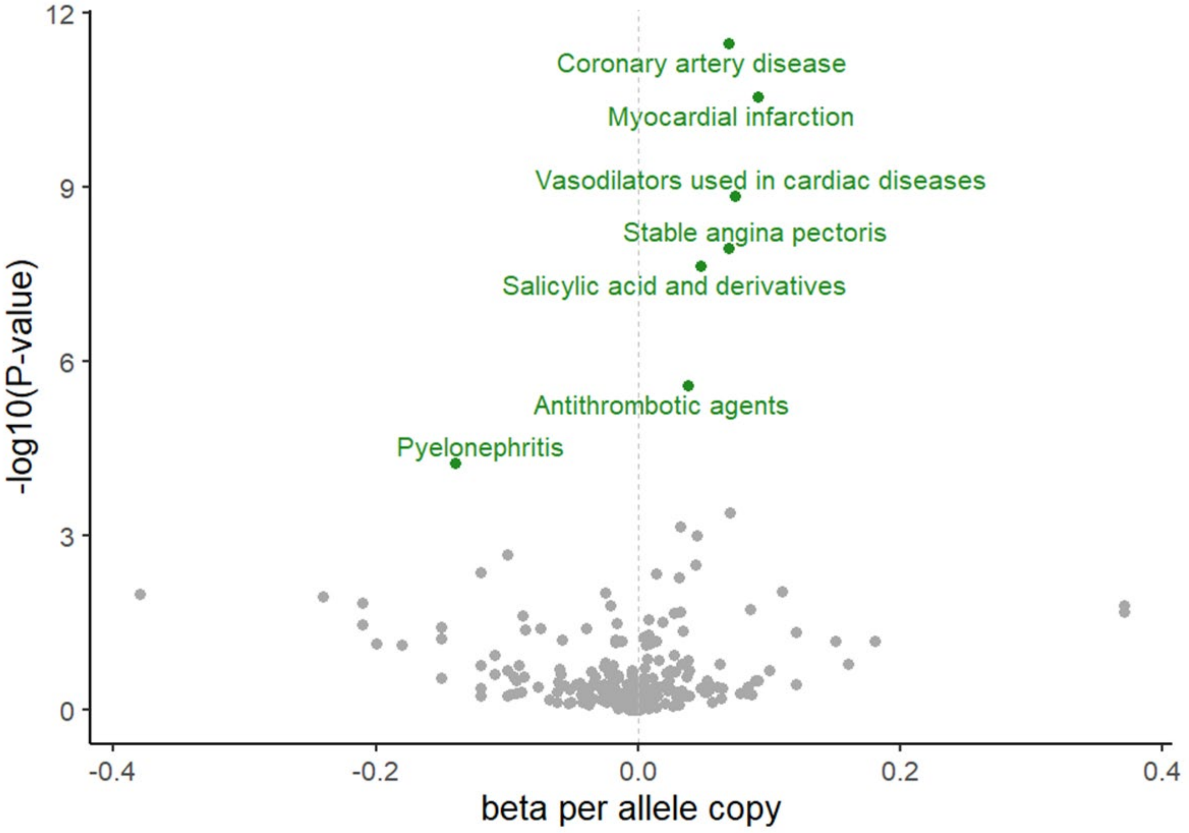


Volcano plot is displaying the association of rs10488763 effect allele T with 259 traits listed in the Biobank Japan PheWeb (https://pheweb.jp/). Vertical grey line indicates no effect. Associations highlighted in green are significant at a P-value threshold of $P=2x10^{-4}$ after correcting for 259 tests.

**Figure 7. Construction of H1-hESC derived cell-lines.**

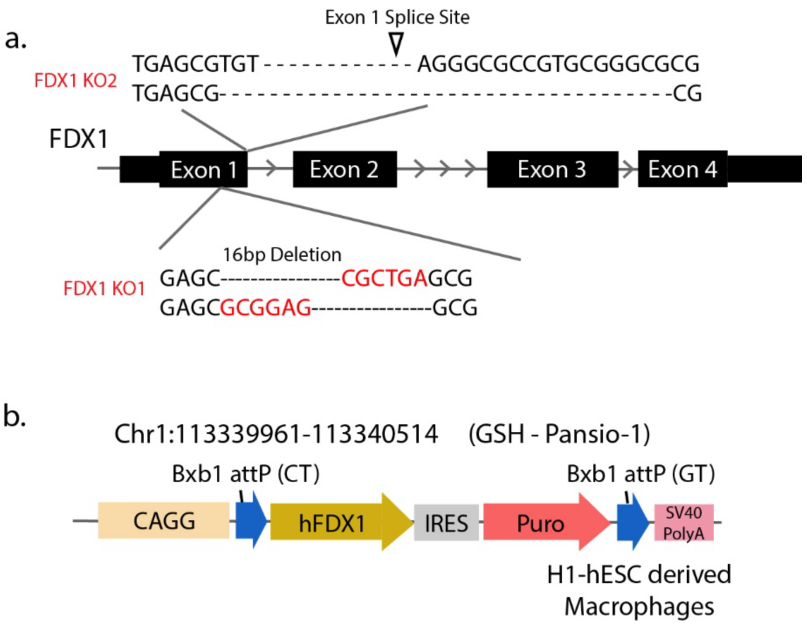


(a) Sequencing validation with two independent FDX1 knock-out (KO) human Embryonic Stem Cell (hESC) clones using two independent sgRNAs targeting Exon 1 splice junction, and a 16-bp deletion on Exon 1 respectively. (b) Schematic illustration of the human FDX1 transgene payload, over-expression (OE) and rescue by knocking into the safe-harbour Pansio-1 locus in H1-hESCs Pansio-1 Safe Harbour line.
